# Oncogene Driver Status Modulates TP53 Prognostic Impact in Advanced NSCLC

**DOI:** 10.64898/2026.09.04.26362258

**Authors:** Minggui Pan, Chen Jiang, Janet Song, Shreyas Kudrimoti, Jacqueline V. Aredo, Heather A. Wakelee, Joel W. Neal, Jenny Zhao, Yasmine Kamgarhaghighi, Ninah S. Achacoso, Aleyda V. Solorzano, Pamela Tse, Elaine Chung, Lori Sakoda, Marie Jennifer Suga, Sachdev Thomas, Laurel Habel

## Abstract

**Purpose:** We compared the association of concurrent TP53 pathogenic variants (PV) with overall survival (OS) by driver status in non-small cell lung cancer (NSCLC).

**Patients and Methods:** The discovery cohort included 3,295 patients with stage IV NSCLC treated at Kaiser Permanent Northern California. The validation cohort included 12,982 NSCLC from 37 public cBioPortal studies. Driver-positive disease was defined as presence of EGFR, ALK, or ROS1 alteration. We used Cox regression modeling to estimate hazard ratios (HRs) for mortality, adjusting for demographics and other factors.

**Results:** Among the discovery cohort, TP53 PV was associated with worse OS among patients with an EGFR, ALK, or ROS1 alteration (HR = 1.80, [95% confidence interval (CI), 1.50-2.15]) but not among those without a driver mutation (HR = 1.08, [95% CI, 0.97-1.21]). Among the validation cohort, TP53 PV was associated with worse OS with HR of 2.27 [1.92-2.69], 2.42 [1.42-4.10], 1.30 [1.20-1.42], and 3.09 [95% CI, 1.47-6.47], for EGFR-activating, ALK-fusion, EGFR/ALK wild-type disease and RET-fusion respectively. The differential association was largest in EGFR exon 20 insertion tumors, in which TP53 PV carried HR = 4.68 [95% CI, 2.34-9.37], 3.9-fold that in driver-negative disease; the corresponding ratio for ERBB2 exon 20 insertions were 1.23 [95% CI, 0.75-2.02]. TP53 PV was not associated with OS in squamous carcinoma.

**Conclusions:** Association of TP53 PV with OS in advanced NSCLC appears to depend on co-occurrence of other driver mutations. This driver-dependent effect modification informs interpretation of TP53 status and may be useful for risk stratification in clinical practice.

## Introduction

TP53 pathogenic variants (PV) are found in approximately half of lung adenocarcinomas and in about four of five squamous cell carcinomas (1,2), making TP53 simultaneously the most common genetic event in non-small cell lung cancer (NSCLC) and one of the least discriminating. A marker present in most tumors cannot easily stratify the disease it defines, and the prognostic literature reflects this: individual series have reported TP53 PV as adverse, as neutral, and occasionally as favorable (3).

Over the same period, treatment of advanced NSCLC has become organized around the presence or absence of a targetable driver alteration. Osimertinib for EGFR-mutated disease, alectinib for ALK-rearranged disease, and crizotinib or entrectinib for ROS1-rearranged disease have each established a survival or progression-free survival advantage (4–7), and KRAS G12C inhibitors have extended targeted therapy into a group previously treated as undruggable (8,9). Prognostic markers are therefore increasingly interpreted within one of these treatment contexts rather than across advanced NSCLC as a whole.

For TP53, one such context is a clear exception. In EGFR-mutated NSCLC treated with tyrosine kinase inhibitors (TKI), concurrent TP53 PV is reproducibly associated with shorter progression-free and overall survival (OS) across many single-institution series and in meta-analysis (3,10–13), with comparable observations in ALK-rearranged disease (14).

What has rarely been asked directly is whether this reflects a genuinely larger TP53 effect in oncogene-driven disease, or simply a better-studied one. The EGFR-mutated literature is almost entirely confined to EGFR-mutated patients, and a recent review states that the magnitude has not been compared against EGFR wild-type (WT) disease (10); one prior study attempted the comparison in 318 patients and found no significant interaction, using a different definition of TP53 PV and considering EGFR alone (16). The comparison is the one with clinical consequence. If TP53 is uniformly adverse, measuring it adds the same information everywhere; if its weight is concentrated in the driver-positive fraction, then TP53 is worth acting on precisely where a targeted therapy decision is already being made, and adds little across the remainder of the disease, which is most of it.

We therefore examined this question in patients with stage IV NSCLC treated within Kaiser Permanente Northern California (KPNC), an integrated healthcare delivery system, with complete demographic and comorbidity capture, unselected referral, and a single sequencing platform, and sought external validation in publicly available data assembled from 483 study archives in cBioPortal. The two data sources have almost opposite deficiencies, so a finding observed in both is unlikely to be an artifact of either.

## Methods

**(additional content in Supplementary Methods)**

***Study setting, population, and data sources***

### Discovery cohort

The discovery cohort was drawn from Kaiser Permanente Northern California (KPNC), an integrated healthcare delivery system serving more than four million members, in which oncology care, pathology, pharmacy dispensing, and vital status are captured within a single electronic health record. We identified 3,295 patients with a histologic diagnosis of stage IV NSCLC who underwent clinical next-generation sequencing (NGS) from November 2017 to December 2023: 2,797 with adenocarcinoma and 498 with squamous cell carcinoma. This group is referred to throughout as the KPNC discovery cohort, and the publicly available data described below as the cBioPortal validation cohort. Small cell carcinoma was excluded by design. Every patient had stage IV disease, so stage is held constant by construction rather than adjusted for, and 97% received first-line systemic therapy. Demographic, comorbidity, and performance status data were drawn from the same record, with comorbidity summarized by the Charlson index (17,18). The study was approved by the KPNC Institutional Review Board with a waiver of informed consent.

OS was measured from the date of diagnosis with delayed entry at the date of NGS, which removes the immortal time between diagnosis and the availability of a genomic result (19,20). This form of left truncation is the principal source of selection bias in survival analyses of genomically profiled cohorts, in which patients are by construction guaranteed to have survived to the date of their own test (21). There were 2,291 deaths, and median OS from the date of NGS was 16.6 months [95% CI, 14.4-19.4] overall, 18.1 months [95% CI, 15.0-21.1] in adenocarcinoma, and 11.1 months [95% CI, 9.7-14.2] in squamous cell carcinoma.

Treatment information in the discovery cohort was limited to whether first-line systemic therapy was received and to the class of that regimen; specific agents, treatment duration, response, and subsequent lines were not extracted, and the validation cohort carries no treatment information at all. The models reported here are therefore not adjusted for therapy.

### Statistical analysis

Prevalence intervals are Wilson score intervals, and between-cohort prevalence differences were tested by Fisher exact test with Benjamini-Hochberg control across 26 comparisons (24). Co-alteration in the validation cohort was tested in whole-exome samples using Cochran-Mantel-Haenszel odds ratios (OR) stratified by within-histology mutation burden quartile (25). Cox regression was used for all survival analyses: in the discovery cohort with delayed entry and 33 to 36 simultaneously entered terms, and in the validation cohort stratified by contributing study. The differential association was quantified as the ratio of the TP53 HR in driver-positive to driver-negative disease, tested by a two-sided z test on the difference of the two log hazard ratios rather than by comparing their significance (26), and in the validation cohort additionally as a multiplicative interaction term. Cohorts were combined by fixed-effect inverse-variance weighting, with heterogeneity tested the same way. All p values are two-sided. The statistical analysis was performed using SAS software version 9.4, R (R Core Team, 2020) and Python 3.14.1.

## Results

### Clinical characteristics by histology and by targetable driver status

Patients with squamous cell carcinoma were four years older (median 73 versus 69 years), more often male (57.2% versus 41.4%), more comorbid (median Charlson index 2 versus 1), and less often Asian (14.5% versus 27.8%) (Supplementary Table S7). Performance status was documented for 9.0% of patients and entered all models with an explicit unknown category. The cohort is stage IV throughout and 97% received first-line systemic therapy, so no comparison below can be generated by stage or by treated-versus-untreated status.

The driver-positive and driver-negative groups differed just as sharply (Supplementary Table S8). Among the 1,053 patients with an EGFR, ALK, or ROS1 alteration, 45.5% were Asian against 16.4% of the driver-negative group, 65.5% were female against 51.9%, and the median age was four years lower. Importantly, TP53 status did not differ between the two groups (53.3% versus 56.5%; Fisher exact p = 0.084), which matters for what follows: the differential association reported below is not a prevalence difference in disguise.

### Determinants of OS in the discovery cohort

In a single Cox model containing 36 terms, worse OS was associated with age, male sex, each Charlson point, ECOG 2-4, and pathogenic variants in TP53 (HR = 1.24, [95% CI, 1.13-1.36]), CDKN2A, SMARCA4, ERBB2, and STK11, together with MYC amplification; better OS with Asian race, driver positivity (HR = 0.74, [95% CI, 0.66-0.84]), and CDK4 PV. KRAS PV was not associated with OS (Figure 1B).

**Figure 1.**
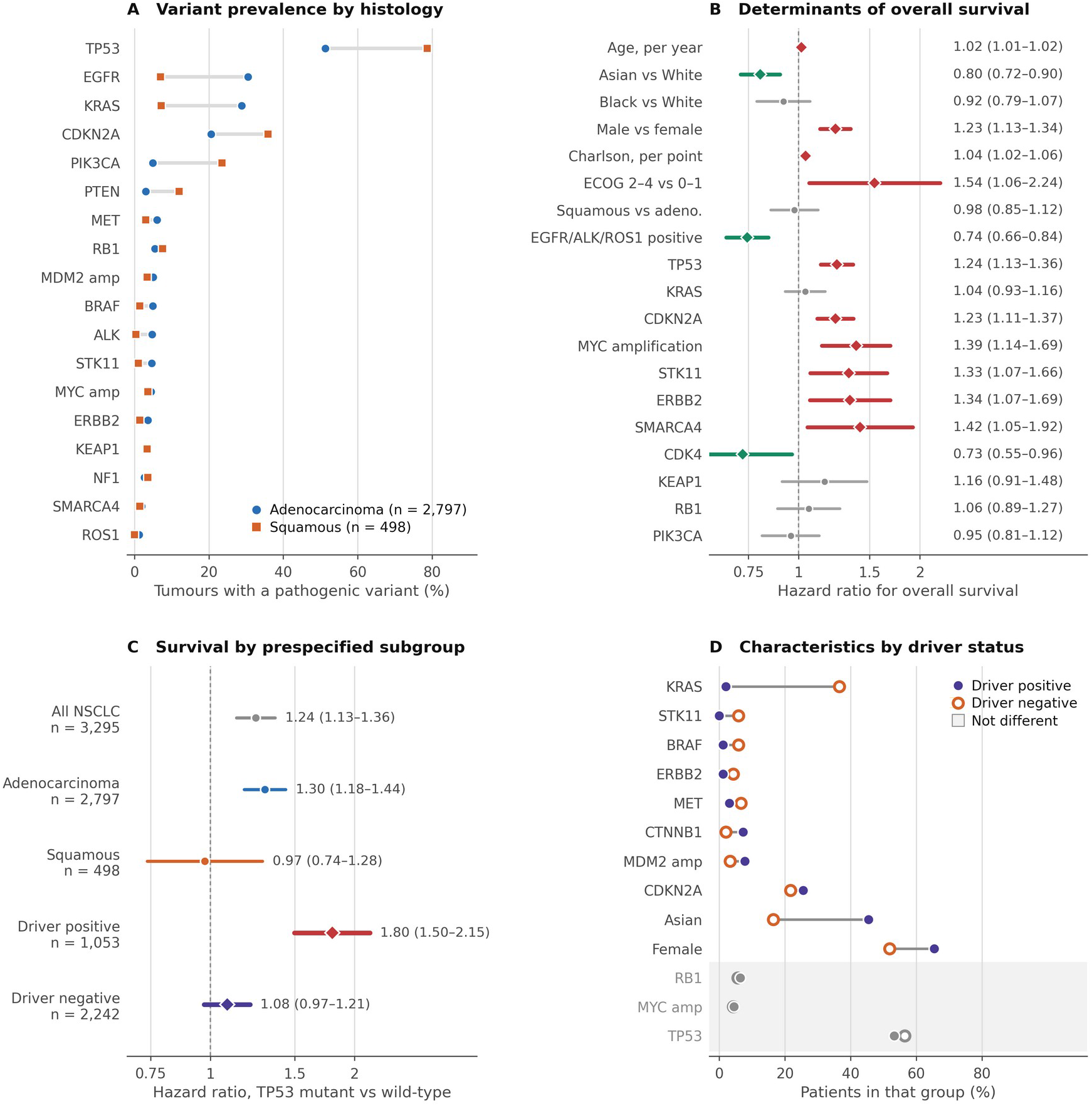
Genomic landscape and determinants of overall survival in the KPNC discovery cohort. (A) Prevalence of pathogenic variants in adenocarcinoma (n = 2,797) and squamous cell carcinoma (n = 498); all differences except those for KEAP1, NF1, RB1, MYC, and SMARCA4 are statistically significant. (B) Hazard ratios for OS for 19 of the 36 terms in the fully adjusted model; red denotes an adverse and green a protective association at p < 0.05, and gray denotes a nonsignificant association. (C) Hazard ratio for TP53 pathogenic variants within each prespecified subgroup; the whole-cohort estimate of 1.24 averages a strong association in driver-positive disease, a weaker one in adenocarcinoma overall, and none in squamous cell carcinoma. (D) Prevalence of selected characteristics in driver-positive (n = 1,053) and driver-negative (n = 2,242) patients; the three shaded rows are the characteristics that do not differ between groups, and TP53 is one of them (53.3% versus 56.5%, p = 0.084). Hazard ratios are from Cox regression models with delayed entry at the date of NGS, adjusted for age, race, sex, Charlson comorbidity index, performance status, histology, and 25 additional genes (n = 3,295; 2,291 deaths). Horizontal bars are 95% confidence intervals. Driver positive denotes a pathogenic EGFR, ALK, or ROS1 alteration.

One finding merits emphasis before the TP53 results. Squamous histology carried an unadjusted HR of 1.36 against adenocarcinoma (p < 0.001), but after adjustment for age, sex, race, comorbidity, performance status, and genotype it was 0.98, [95% CI, 0.85-1.12].

### Differential OS association of TP53 PV by targetable driver status among discovery cohort

The whole-cohort TP53 estimate of HR = 1.24 comprises three different quantities (Figure 1C). Within adenocarcinoma it was 1.30, [95% CI, 1.18-1.44]; within squamous cell carcinoma 0.97, [95% CI, 0.74-1.28], no signal at all in a histology in which 78.7% of tumors carry a TP53 PV. Within the driver-positive group it was 1.80, [95% CI, 1.50-2.15] and within the driver-negative group 1.08, [95% CI, 0.97-1.21], so among the two thirds of patients without a targetable driver TP53 carried no detectable prognostic information. Expressed as a ratio of hazard ratios, the driver-positive estimate was 1.66 times the driver-negative one [95% CI, 1.34-2.05], p < 0.001.

### Validation of the differential association in an independent cohort

The validation cohort was assembled from public data with no reference to the discovery results; it is 3.9-fold larger in adenocarcinoma and resolves every mutation to an allele, but has no comorbidity, performance status, or treatment information, and 63% derives from a single institution’s prospective sequencing program (27) (Supplementary Table S1, Supplementary Figure S1). Within validation adenocarcinoma, TP53 alteration carried HR = 2.27, [95% CI, 1.92-2.69] in EGFR-activating tumors, HR = 2.42, [95% CI, 1.42-4.10] in ALK-fusion tumors, and HR = 1.30, [95% CI, 1.20-1.42] in EGFR/ALK-WT tumors (Supplementary Table S9); mutual adjustment for 15 and 17 additional genes left the contrast intact (2.24 against 1.32). In squamous cell carcinoma TP53 yielded HR = 0.97 and 0.93 before and after mutual adjustment. The driver-negative estimate reached significance here and not in the discovery cohort, which a cohort 2.4-fold larger should produce; the point estimates are of the same modest size.

Expressed as the ratio of the TP53 PV HR in driver-positive to that in driver-negative disease, the discovery cohort yielded 1.66 [95% CI, 1.34-2.05] and the validation cohort 1.74 [95% CI, 1.44-2.10], with no detectable heterogeneity (p = 0.74) and a pooled estimate of 1.70 [95% CI, 1.48-1.96], p < 0.001 (Figure 2, Supplementary Table S2). Estimated instead as an explicit multiplicative interaction term within a single validation model, the same quantity was 1.67 [95% CI, 1.38-2.01].

**Figure 2.**
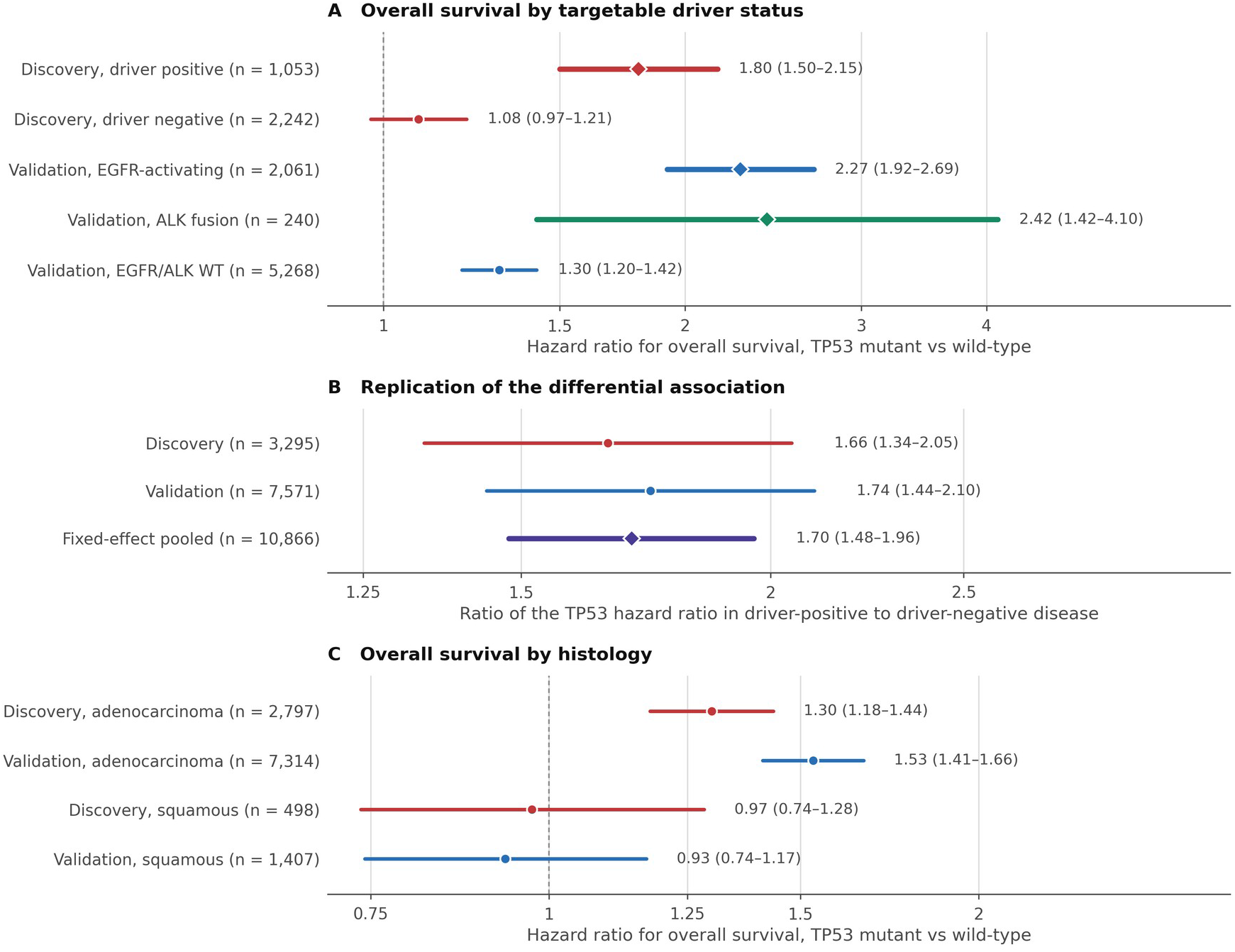
Replication of the differential association in the cBioPortal validation cohort. (A) Hazard ratio for TP53 pathogenic variants within each driver stratum in both cohorts. (B) Ratio of the TP53 hazard ratio in driver-positive to driver-negative disease, with the fixed-effect pooled estimate; a ratio of 1 would indicate that TP53 carries equal prognostic weight with and without a targetable driver. (C) Hazard ratio for TP53 pathogenic variants by histology in both cohorts; between-cohort heterogeneity was p = 0.81 for squamous cell carcinoma and p = 0.013 for adenocarcinoma. Discovery estimates are adjusted for age, race, sex, comorbidity, performance status, histology, and 25 additional genes; validation estimates are stratified by contributing study. Horizontal bars are 95% confidence intervals and n denotes the patients contributing to each survival model. Between-cohort heterogeneity was tested by a two-sided z test on the difference of log hazard ratios.

TP53 PV was not associated with OS in squamous cell carcinoma in either cohort. The comparably adjusted estimates of HR = 0.97, [95% CI, 0.74-1.28] and HR = 0.93, [95% CI, 0.74-1.17] differ by less than 5% (p for heterogeneity = 0.81), in a histology in which TP53 is altered in 78.7% and 84.6% of tumors respectively. In adenocarcinoma overall the two cohorts yielded HR = 1.30 and HR = 1.53, formally heterogeneous (p = 0.013).

The validation cohort also reproduced the demographic signature of driver-positive lung cancer. Against EGFR/ALK-WT adenocarcinoma, EGFR-activating tumors were more often female, more often Asian, and four years younger, and ALK-fusion tumors ten years younger (all p < 0.001), matching the discovery contrasts in direction and approximate magnitude.

### Smoking does not account for the differential association

The validation cohort documented smoking history for 32.3% of patients with adenocarcinoma, including 3,120 with survival data. Never-smoker enrichment was marked: 58.1% of EGFR-activating and 69.2% of ALK-fusion tumors arose in never smokers against 18.5% of EGFR/ALK-WT tumors (OR = 6.12 and 9.93, both p < 0.001), consistent with the recognized distinctiveness of lung cancer in never smokers (28,29).

Among those 3,120 patients the TP53 PV by driver interaction was 1.661 [95% CI, 1.228-2.247] without a smoking term and 1.658 with one, a change of 0.2%; adding age and sex gave 1.72, and adjusting for stage in a separate subset of 3,332 patients gave 1.57 [95% CI, 1.18-2.08]. Stratification gave the same answer directly: among never smokers the ratio was 1.72 [95% CI, 1.05-2.83], p = 0.031, and among ever smokers 1.39 [95% CI, 0.90-2.15], the direction being consistent in both strata (Supplementary Figure S2, Supplementary Table S3).

### TP53 alteration frequency and mutation spectrum by driver group

TP53 alteration reached 52.5% [95% CI, 50.7-54.2] in EGFR-activating and 25.7% [95% CI, 21.6-30.3] in ALK-fusion tumors against 46.0% [95% CI, 44.9-47.2] in WT tumors (both p < 0.001). ALK-fusion adenocarcinoma was the least TP53-altered lung group in the atlas (Figure 3A) (15,30).

**Figure 3.**
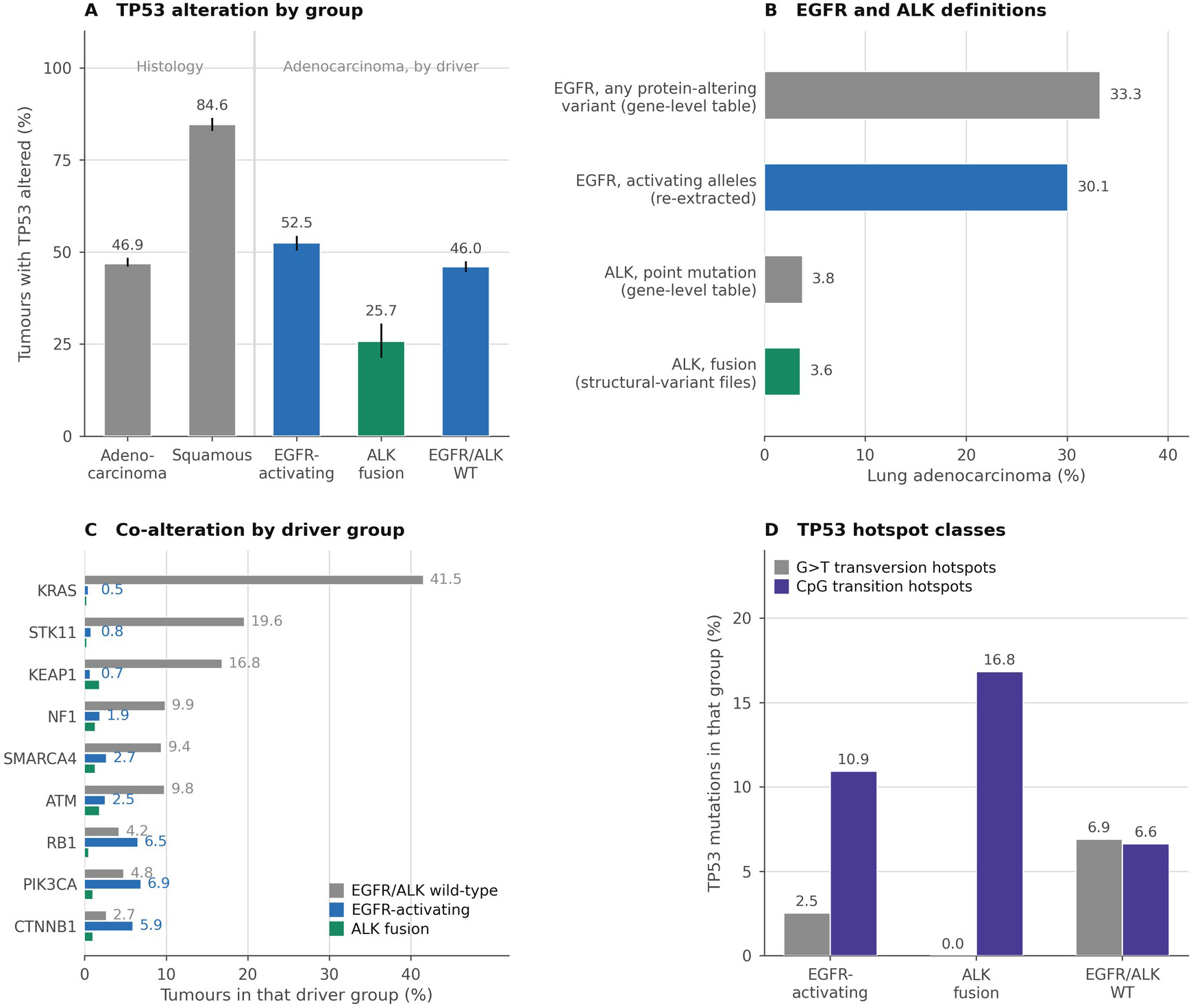
Analyses requiring allele-and fusion-level resolution in the cBioPortal validation cohort. (A) TP53 alteration by histology and, within adenocarcinoma, by driver group; vertical bars are Wilson 95% confidence intervals. ALK-fusion adenocarcinoma is the least TP53-altered group (25.7% versus 46.0% in wild-type; OR = 0.41, p < 0.001) whereas EGFR-activating tumors are slightly more altered (52.5%; OR = 1.29, p < 0.001). (B) Prevalence of EGFR and ALK alterations under gene-level and under allele– or fusion-resolved definitions; the gene-level EGFR call overstates the targetable group by 11%, and the two ALK calls are of similar size but identify different tumors, with only 35 carrying both. (C) Prevalence of co-altered genes within each driver group; values are printed for the wild-type and EGFR-activating groups. (D) Proportion of TP53 mutations occurring at G>T transversion and CpG transition hotspots, by driver group. Validation cohort only (n = 10,797 adenocarcinoma; 2,185 squamous cell carcinoma). Co-alteration was tested in whole-exome samples by Cochran-Mantel-Haenszel stratification on mutation burden quartile.

Tobacco-associated G>T transversion hotspots comprised 6.9% of TP53 mutations in WT adenocarcinoma, 2.5% in EGFR-activating, and 0.0% in ALK-fusion tumors, whereas CpG transition hotspots ran 6.6%, 10.9%, and 16.8% (EGFR versus WT: transversion OR = 0.35; CpG OR = 1.73, both p < 0.001). The leading variants make the point without statistics: V157F, R158L, and R273L, all lung-enriched smoking-associated alleles (31–33), against R273C, R273H, and R248W in EGFR-activating tumors. Co-alteration differed markedly (Figure 3C): KRAS PV was present in 41.5% of WT adenocarcinoma against 0.46% of EGFR-activating tumors, STK11 in 19.6% against 0.80%, and KEAP1 in 16.8% against 0.68% (34–36). RB1 was the exception, co-occurring with TP53 in both groups (37).

### Concordance between the two cohorts and the effect of variant interpretation

We estimated 16 gene-level survival associations in both cohorts, of which 13 were statistically consistent (Supplementary Table S4). Agreement was closest for the largest effects: MYC amplification 1.45 against 1.47, SMARCA4 1.38 against 1.42, STK11 1.24 against 1.31, KEAP1 1.31 against 1.46. Driver positivity was protective in both and KRAS null in both. Of the three exceptions, TP53 in adenocarcinoma differed in degree as described above, CDKN2A because the discovery call combines mutation and deletion, and STK11 in squamous cell carcinoma rests on five mutated patients.

Among the 12 oncogene comparisons the median discovery-to-validation prevalence ratio was 1.00, whereas among the 12 tumor suppressor comparisons it was 0.38 (STK11 4.6% against 13.2%, KEAP1 3.1% against 11.4%, SMARCA4 1.9% against 7.1%) (22,23,38). These genes nonetheless carried almost identical hazard ratios (Figure 4).

**Figure 4.**
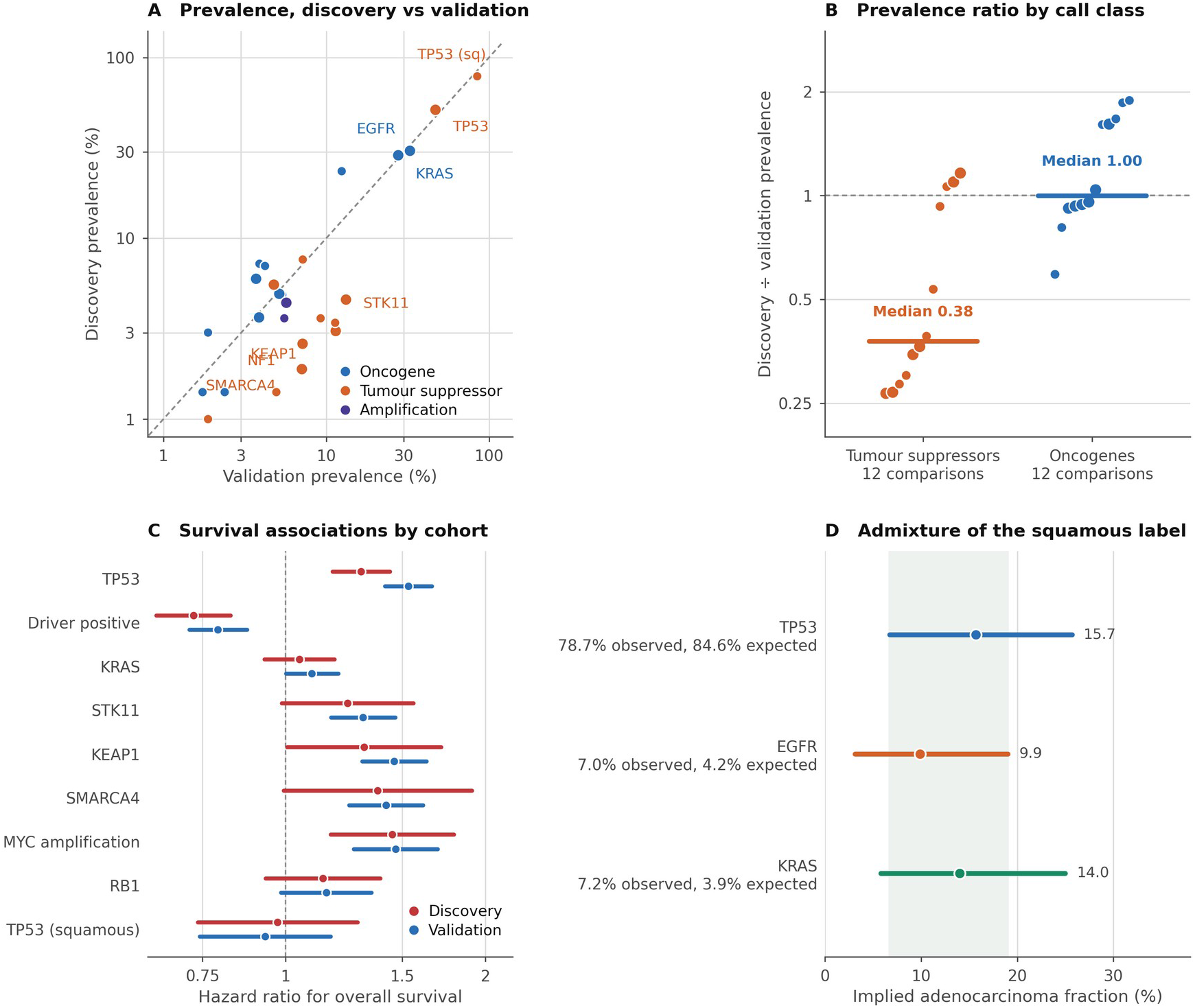
Concordance between the two cohorts and its determinants. (A) Prevalence of each gene in the discovery against the validation cohort; the dashed line denotes equality, large markers denote adenocarcinoma and small markers squamous cell carcinoma. Oncogenes lie on the line whereas STK11, KEAP1, NF1, and SMARCA4 lie approximately threefold below it. (B) Ratio of discovery to validation prevalence by variant-call class, with group medians; the two tumor suppressors near 1 are TP53 and RB1, whose variant spectra are already exhaustively catalogued. (C) Hazard ratios for nine of the 16 survival associations in Supplementary Table S4, selected for legibility, estimated separately in each cohort; 13 of the 16 are statistically consistent. (D) Fraction of the discovery squamous group implied to be adenocarcinoma, estimated separately from three markers; shading denotes the interval common to all three. This is an audit heuristic rather than a calibrated method and does not affect any adenocarcinoma result. Prevalence differences were tested by Fisher exact test with Benjamini-Hochberg control across 26 comparisons, and survival differences by a two-sided z test on the difference of log hazard ratios.

### RET, HER2, NTRK, and NRG1 in the cBioPortal validation cohort

The four remaining targetable drivers were assessed directly in the validation cohort. RET fusion was present in 1.67% of adenocarcinomas [95% CI, 1.42-1.95], ERBB2 activating mutation in 2.65% [95% CI, 2.36-2.97], NTRK1/2/3 fusion in 0.24%, and NRG1 fusion in 0.20% (Supplementary Table S10, Figure 5A); all four were essentially absent from squamous cell carcinoma. Gene-level calls misled for both: they would make RET a frequent squamous driver when the converse is true, and overstated the ERBB2 activating group by 46% against 11% for EGFR (Figure 5B).

**Figure 5.**
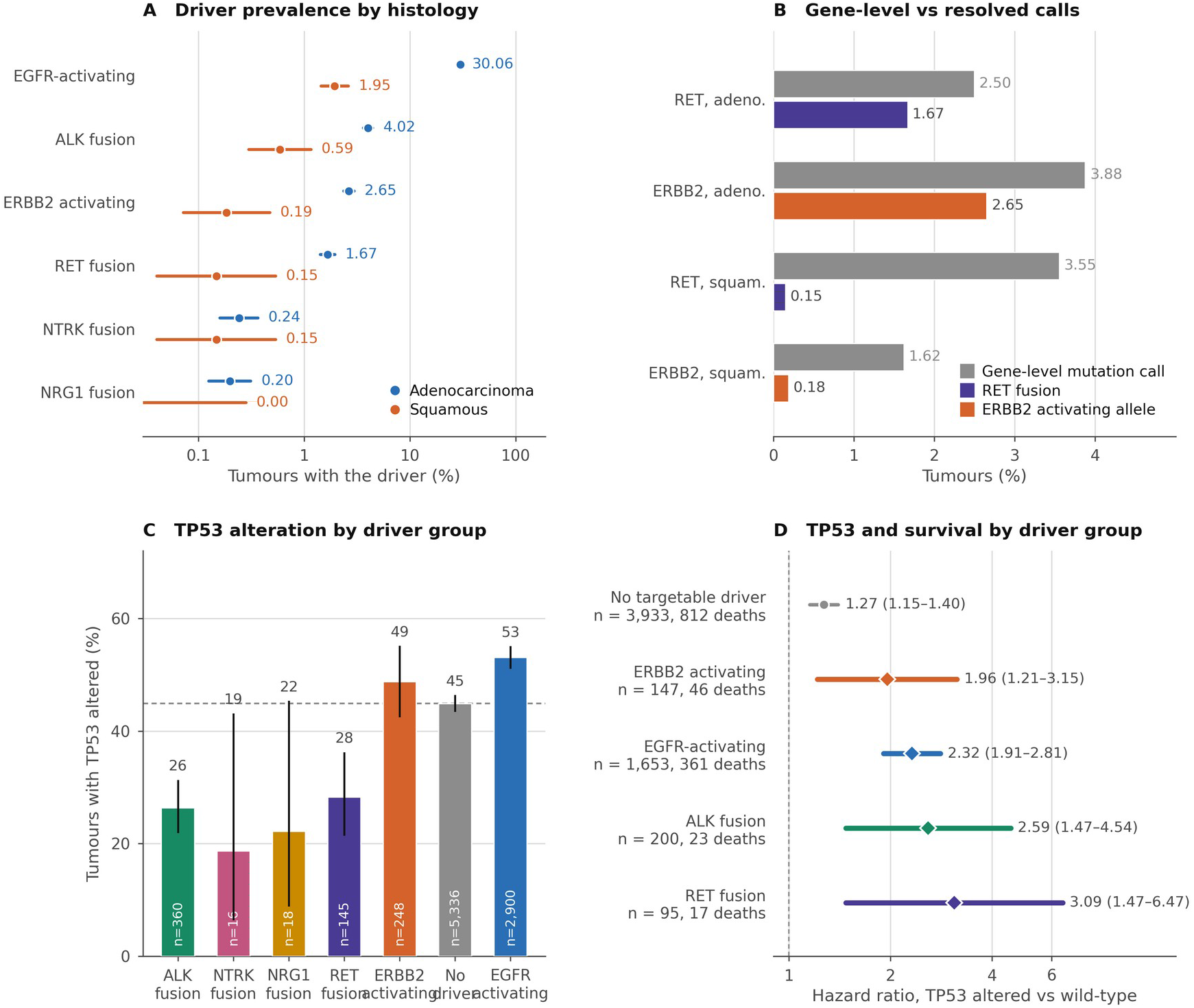
RET, HER2, NTRK, and NRG1 in the cBioPortal validation cohort. (A) Prevalence of each targetable driver in adenocarcinoma and in squamous cell carcinoma, with Wilson 95% confidence intervals; note the logarithmic scale. (B) Prevalence under a gene-level mutation call against the event-resolved call, computed on the same denominator within each row; for RET the two calls overlap in only 4 adenocarcinomas and in no squamous carcinoma. (C) TP53 alteration by driver group, with Wilson 95% confidence intervals; the dashed line is the value in the absence of a targetable driver. (D) Hazard ratio for TP53 alteration within each driver group, from Cox regression models stratified by contributing study. Fusion prevalence is computed among the 9,052 adenocarcinomas from studies depositing structural variants and ERBB2 prevalence among all 10,788. NTRK and NRG1 fusions were too infrequent to support a survival model.

TP53 alteration segregated by driver class rather than by targetability (Figure 5C). Against 44.9% without a targetable driver, TP53 was altered in 28.3% of RET-fusion tumors (OR = 0.48, q < 0.001), 26.4% of ALK-fusion tumors (OR = 0.44, q < 0.001), and 18.8% and 22.2% of NTRK-and NRG1-fusion tumors, against 53.1% of EGFR-activating tumors (OR = 1.39, q < 0.001). ERBB2-activating tumors were the exception at 48.8% (OR = 1.17, q = 0.28).

Among 6,069 patients with survival data, RET fusion, EGFR-activating, and ALK-fusion disease were each associated with better OS, whereas ERBB2-activating disease was not (Supplementary Table S10), the opposite of the ordering reported in stage I-III disease (39). Within each group, TP53 alteration carried HR = 3.09 [95% CI, 1.47-6.47] in RET-fusion, 2.59 [95% CI, 1.47-4.54] in ALK-fusion, 2.32 [95% CI, 1.91-2.81] in EGFR-activating, and 1.96 [95% CI, 1.21-3.15] in ERBB2-activating tumors, against 1.27 [95% CI, 1.15-1.40] without a targetable driver (Figure 5D), giving ratios of 2.78, 1.93, 1.81, and 1.34. The RET and ALK estimates rest on 17 and 23 deaths among TP53-altered patients, so the ordering is provisional; the RET estimate is consistent with a series of 129 patients with RET-rearranged NSCLC in which TP53 mutation carried HR = 2.26 (40). Substituting the six-driver definition left the overall ratio unchanged at 1.75 (Supplementary Table S5).

### Exon 20 insertion classes in both cohorts

In the validation cohort, ERBB2 exon 20 insertions accounted for most activating HER2 alterations, 223 of 10,788 adenocarcinomas (2.07% [95% CI, 1.82-2.35]) against 63 with another activating allele, and were almost confined to adenocarcinoma; three alleles covered 92.8% of the group (Figure 6A).

**Figure 6.**
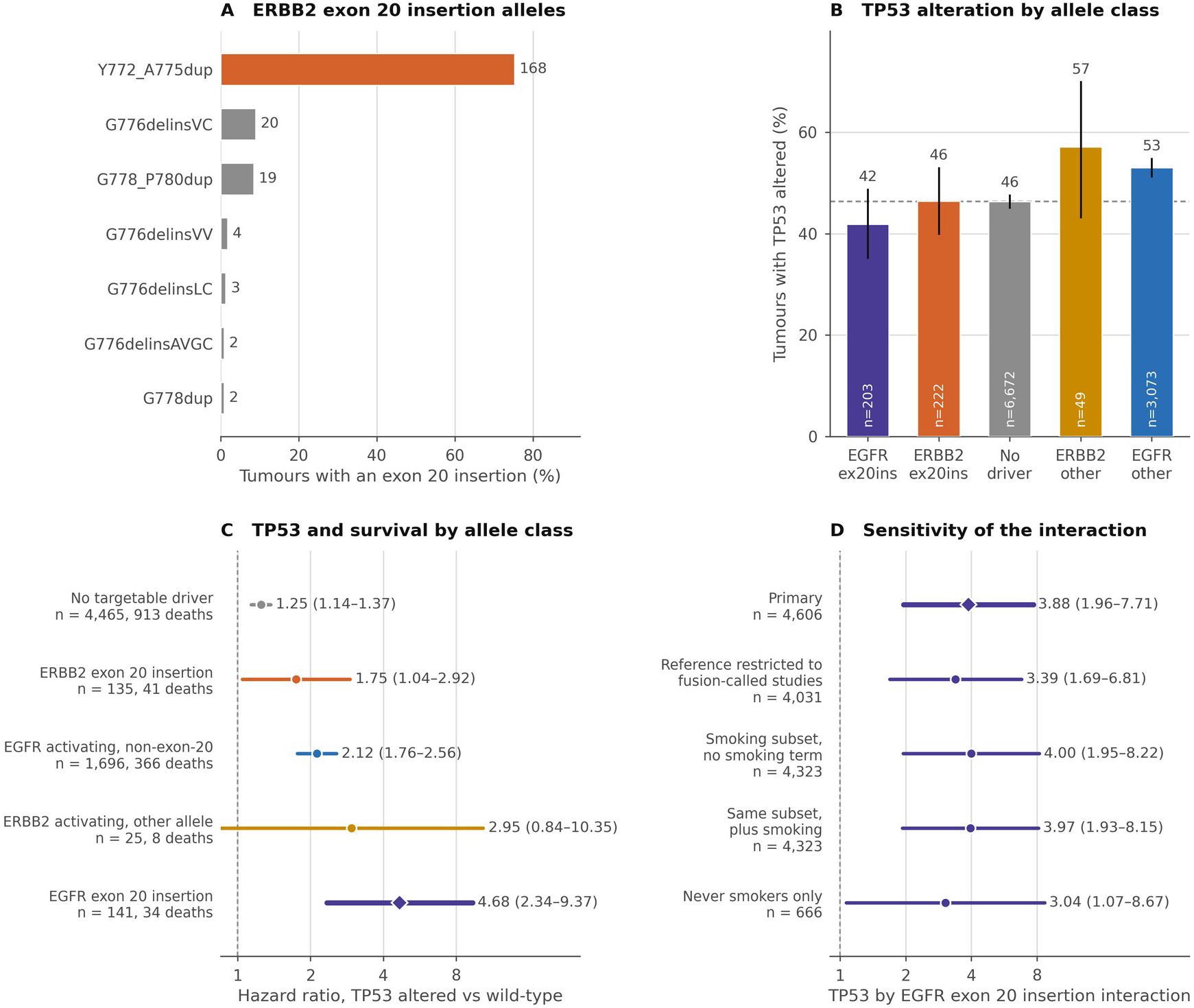
ERBB2 and EGFR exon 20 insertions in the cBioPortal validation cohort. (A) Allele distribution among the 223 adenocarcinomas carrying an ERBB2 exon 20 insertion; the seven commonest alleles are shown and the bar labels are tumor counts. (B) TP53 alteration by allele class, with Wilson 95% confidence intervals; the dashed line is the value in the absence of a targetable driver. (C) Hazard ratio for TP53 alteration within each allele class, from Cox regression models stratified by contributing study. (D) Sensitivity of the TP53 by EGFR exon 20 insertion interaction to the choice of reference group and to smoking. Horizontal bars are 95% confidence intervals.

TP53 alteration in ERBB2 exon 20 insertion tumors was 46.4% [95% CI, 40.0-53.0], indistinguishable from the 46.4% seen without a targetable driver (OR = 1.00, [95% CI, 0.77-1.31]). Mutual exclusivity with KRAS was absolute, and STK11 and KEAP1 were infrequent at 2.7% and 1.4% against 20.9% and 17.9%.

Separating the EGFR exon 20 insertion from the other activating EGFR alleles showed that the two insertion classes did not differ from each other in TP53 alteration frequency (OR = 1.20, p = 0.38), whereas both differed from the alleles the EGFR insertion is usually pooled with: 41.9% against 53.0% (OR = 0.64, p = 0.002) (Figure 6B).

The survival findings differed between the two classes. Among 6,798 validation patients, neither ERBB2 exon 20 insertion (HR = 1.23, [95% CI, 0.96-1.57]) nor EGFR exon 20 insertion (HR = 0.79, [95% CI, 0.59-1.06]) differed from the absence of a targetable driver, against HR = 0.67, [95% CI, 0.61-0.74] for the other activating EGFR alleles. Within groups, TP53 alteration carried HR = 1.75, [95% CI, 1.05-2.92] in ERBB2 exon 20 insertion tumors and HR = 4.68, [95% CI, 2.34-9.37] in EGFR exon 20 insertion tumors, against 2.12 in other activating EGFR tumors and 1.25 without a targetable driver (Supplementary Table S11, Figure 6C); the ratios against driver-negative disease were 3.88 [95% CI, 1.96-7.71] and 1.23 [95% CI, 0.75-2.02]. The EGFR estimate is the largest effect modification in the validation cohort and rests on 141 patients (Figure 6D, Supplementary Table S6).

The same separation was applied within the KPNC discovery cohort. Of 109 patients with a pathogenic ERBB2 variant, 60 (55.0%) carried an exon 20 insertion, and of 887 with a pathogenic EGFR variant, 51 (5.7%) did, close to the 6.2% seen in validation. TP53 PV was less frequent in both insertion classes than in the alleles with which they are usually pooled, 51.7% against 73.5% for ERBB2 (p = 0.020) and 45.1% against 59.0% for EGFR (p = 0.051), reproducing the validation direction and magnitude. Survival in neither class was associated with TP53 PV: unadjusted HR = 0.87, [95% CI, 0.48-1.57], p = 0.65 for ERBB2 exon 20 insertion on 47 deaths, and HR = 1.25, [95% CI, 0.68-2.30], p = 0.47 for EGFR exon 20 insertion on 43 deaths (Supplementary Table S12).

## Discussion

In 3,295 patients with stage IV NSCLC from an integrated healthcare organization and an independent validation cohort of 12,982 tumors, the prognostic association of TP53 PV with OS differed according to targetable driver status. With full demographic, comorbidity, and performance status adjustment, TP53 PV was adverse among patients with a targetable driver (HR = 1.80) but not among those without one (HR = 1.08). Public data reproduced the contrast using allele– and fusion-resolved groups, the two estimates of the differential association being 1.66 and 1.74. The prognostic weight of TP53 in advanced NSCLC therefore appears to be a property of tumor biology rather than of the gene itself.

The adverse effect of concurrent TP53 PV in EGFR-mutated disease is among the better replicated observations in thoracic oncology (3,10,11); what is new here is the comparison, which a recent review notes has not been made against EGFR-WT disease (10). The two halves have been reported separately, an adjusted HR of 2.62 [95% CI, 1.98-3.46] among 436 patients with a rare treatable driver (41) and no association among 762 patients with driver-negative disease (adjusted HR = 1.15 [95% CI, 0.96-1.38]) (42), but neither estimated the contrast within a single population; that analysis also found the association to depend on TP53 mutation class rather than binary status (42), consistent with the different mutational processes evident in our two driver groups. The closest prior attempt found nondisruptive TP53 mutations adverse in both EGFR strata of 318 patients without comparing them, at a sample size whose interval would span our estimate of 1.70 and the null alike (16).

Driver-positive patients receive TKI and driver-negative patients chemotherapy or immunotherapy, and given the benefit demonstrated for driver-directed therapy in randomized trials (4–7), the survival advantage of the driver-positive group (HR = 0.73 and 0.79) is in large part a treatment effect. TP53 may therefore be prognostic in driver-positive disease because it predicts the duration of TKI benefit, a pharmacodynamic rather than an intrinsic property, consistent with reports that TP53 co-mutation accelerates the evolution of resistance in EGFR-mutated disease (14,43).

The exon 20 insertion findings constrain this account, and the two cohorts do not agree. In validation the TP53 PV association was more than twice as large in EGFR exon 20 insertion tumors as in those carrying a classical activating allele (HR = 4.68 versus 2.12), whereas HER2 exon 20 insertion carried the smallest ratio against driver-negative disease of any targetable group (1.23), although the association within that group was itself significant (HR = 1.75) and a prior series of 112 ERBB2 exon 20 insertion tumors reported concurrent TP53 mutation to be adverse (44). The discovery cohort reproduces neither, returning 1.25 and 0.87, and the EGFR difference is significant (p = 0.005). It is explained neither by class definition, since restricting the discovery class to in-frame insertions matched the validation prevalence without moving the hazard ratio, nor by differential access to matched therapy, since both classes lacked an effective inhibitor for most of the accrual period. What both cohorts show is that TP53 carries no more prognostic weight in HER2 exon 20 insertion disease than in disease without a targetable driver. The validation EGFR exon 20 estimate therefore requires confirmation the discovery cohort does not supply.

TP53-mutant NSCLC shows higher tumor mutational burden, higher PD-L1 expression, and a more inflamed, T cell-infiltrated microenvironment, and TP53 mutation has been associated with better response and longer survival on immune checkpoint blockade, most strongly in KRAS-mutated adenocarcinoma (45–47). Our driver-negative group fits that description and received checkpoint inhibitors as standard first-line therapy across the accrual period, whereas driver-positive patients receive a TKI. If TP53 is adverse for intrinsic tumor biology but favorable for immunotherapy sensitivity, the two effects would partially cancel in driver-negative disease and not in driver-positive disease, attenuating the observed association in the former.

The clinical corollary is under direct test. In a randomized trial reported in 2026, 294 patients with untreated advanced EGFR-mutated non-squamous NSCLC and a concurrent TP53 mutation received osimertinib with or without carboplatin and pemetrexed; median progression-free survival was 34.0 against 15.6 months (HR = 0.44, [95% CI, 0.32-0.61]), with an early overall survival signal (HR = 0.57, [95% CI, 0.37-0.87]) (48). That trial used TP53 status as the entry criterion for treatment intensification rather than as a prognostic descriptor, the strongest available demonstration that TP53 is clinically actionable in driver-positive disease.

The discovery cohort’s strengths are unselected referral within an integrated healthcare organization, a single sequencing platform, essentially complete capture of demographics, comorbidity, and vital status, and stage and treatment status held constant by design; delayed entry at the date of NGS removes the immortal time that inflates survival estimates in sequenced cohorts. The validation cohort is nearly fourfold larger, resolves EGFR to the allele and ALK to the fusion, and records smoking, so the two datasets have largely complementary limitations and a finding present in both is unlikely to be an artifact of either.

Our study also has limitations. The class of first-line therapy is confounded with driver status by definition and was not entered into the models, and the validation cohort carries no treatment data, so we cannot yet distinguish a differential biological effect of TP53 from a differential treatment effect. The discovery exon 20 analyses are unadjusted and rest on 51 and 60 patients. Patients with a KRAS G12C, MET exon 14 skipping, or BRAF V600E alteration were retained in the driver-negative group, and the few driver-negative patients carrying a RET, HER2, NTRK, or NRG1 alteration are biologically closer to the driver-positive group, both of which bias the contrast toward the null. The findings are restricted to stage IV disease (49); the earlier immunohistochemical literature in resected disease reported the opposite histologic ordering (50) in a different assay, stage, and era.

In summary, the prognostic association of TP53 PV with OS in advanced NSCLC is concentrated in tumors carrying a targetable driver alteration. This pattern was consistent across two cohorts with complementary methodological weaknesses and was not explained by stage, by treatment receipt, or by smoking. TP53 status therefore carries clinically meaningful information precisely where a targeted therapy decision is being made.

## Corresponding Author

Minggui Pan, MD, PhD, Division of Oncology, Stanford University School of Medicine, Stanford, CA 94305.

## Data Availability

All validation source data are publicly available from cBioPortal. The derived per-sample lung datasets, the re-extracted EGFR and ALK oncogene and fusion records, the re-extracted patient demographics and smoking history, and the full tables underlying every figure accompany this manuscript as supplementary data files. Data from the discovery cohort are governed by the sponsoring healthcare organization and are not publicly redistributable; the summary statistics required to reproduce every comparison reported here are provided in the accompanying tables.

## AI disclosure

The authors acknowledge the use of Claude Cowork and Gemini 3, developed by Anthropic and Google, to assist in drafting and optimizing the Python scripts used for survival analyses described in the Methods section. All code, data outputs, and interpretations generated with this assistance were independently cross-checked, validated, and verified by the human authors. The authors retain absolute accountability for the scientific accuracy of the data and the conclusions drawn.

## Supplementary Methods

### NGS and definition of pathogenic variants

All discovery-cohort sequencing was performed on StrataNGS (Strata Oncology, Ann Arbor, MI), a CLIA-certified, CAP-accredited comprehensive genomic profiling test using multiplex PCR and semiconductor sequencing on DNA and RNA co-isolated from the same formalin-fixed, paraffin-embedded specimen (S1). The 429-gene panel reports single-nucleotide variants, short indels, copy number alterations, and RNA-based gene fusions, so fusion drivers including ALK, ROS1, RET, NTRK1/2/3, and NRG1 are detectable on the same assay. A single assay in a single reference laboratory was used for every patient across the accrual period. Genomic calls are the clinical PV report, made under established professional-society frameworks (22,23); a tumor was counted as altered if the report listed a pathogenic or likely pathogenic variant, with copy number gains reported separately as amplification. TP53 was modeled as binary throughout.

### Definition of targetable driver status in the discovery cohort

Ten oncogenic drivers in NSCLC now have at least one approved matched targeted therapy: EGFR, ALK, ROS1, RET, KRAS G12C, MET exon 14 skipping, HER2 (ERBB2), BRAF V600E, NTRK fusions, and NRG1 fusions. We defined driver-positive disease as a clinically reported pathogenic EGFR, ALK, or ROS1 alteration, comprising 1,053 patients against 2,242 without. These three were chosen because matched first-line tyrosine kinase inhibitors for EGFR-, ALK-, and ROS1-altered NSCLC were guideline-endorsed standard of care throughout the entire accrual window, so the driver-positive group is internally uniform with respect to treatment availability, whereas approvals for the remaining seven drivers fell during or after that window; and because these three define a group large enough to support the stratified survival modeling reported here.

Patients whose tumors carried a KRAS G12C mutation, a MET exon 14 skipping alteration, or a BRAF V600E mutation were deliberately retained in the driver-negative group, which is stated explicitly because such tumors are not driver-negative in the strict biological sense. They were retained because matched inhibitors were unavailable or restricted to later lines for most of the accrual window, so nearly all received platinum-based chemotherapy with or without an immune checkpoint inhibitor first line, as the remainder of the driver-negative group did (Supplementary Methods). Driver-negative should therefore be read throughout as the absence of an EGFR, ALK, or ROS1 alteration rather than of any targetable oncogenic driver.

ERBB2 and EGFR pathogenic variants in the discovery cohort were also separated into an exon 20 insertion class, admitting in-frame insertions only, and a non-exon-20 class carrying T790M and other non-insertion exon 20 variants; survival within each insertion class was modeled with TP53 as the only term (Supplementary Methods).

### Validation cohort

We accessed 483 study archives from cBioPortal (S2,S3) and constructed a non-redundant cohort of 209,672 unique samples, excluding cell lines, xenografts, and liquid biopsies. The NSCLC subset comprised 10,797 adenocarcinomas and 2,185 squamous cell carcinomas from 37 studies, classified on the OncoTree-derived detailed cancer type (S5); small cell carcinoma was excluded to match the discovery cohort. OS was available for 7,571 patients with adenocarcinoma (2,828 deaths) and 1,446 with squamous cell carcinoma. Because 37 studies of differing design, follow-up, and treatment era contribute, and the ten largest supply 92% of the tumors, every survival model in this cohort was stratified by contributing study. TP53 mutation was defined as any missense, nonsense, frameshift, canonical splice, in-frame indel, nonstop, or translation start site variant, and TP53 alteration additionally included GISTIC-2 deep deletion (S6). Cohort assembly, demographic re-extraction, and vocabulary harmonization are detailed in the Supplementary Methods.

### Definition of oncogene-driven disease in the validation cohort

The gene-level driver table used for most public analyses is inadequate for defining oncogene-driven lung cancer. For EGFR the problem is quantitative: counting any protein-altering variant gives 33.3% of adenocarcinoma against 30.1% for activating alleles, overstating the targetable group by 11%. For ALK it is categorical: point mutations and fusions occur at similar frequency but identify different tumors, and the disease-defining event is the EML4-ALK fusion (S7), which resides in structural-variant files. We therefore re-extracted oncogene protein-level and fusion records from all 37 lung-contributing archives, yielding three mutually exclusive adenocarcinoma groups: 3,239 EGFR-activating, 385 ALK-fusion, and 7,160 EGFR/ALK-WT tumors. Only 19 of 37 archives deposited structural variants, so ALK-fusion prevalence is a lower bound. RET, NTRK1/2/3, and NRG1 fusions and ERBB2 activating alleles were called on the same principle, and the ERBB2 and EGFR exon 20 insertion classes were separated from the other activating alleles of each gene so the two structurally analogous insertions could be compared. The allele and fusion definitions, the reconciliation of tumor counts, and the one-row-per-patient survival convention are given in full in the Supplementary Methods. The two driver definitions are therefore not identical, the discovery one being a gene-level clinical call including ROS1 and the validation one an allele– and fusion-resolved research call excluding it.

### TP53 and covariate definitions in the validation cohort

For the mutational process analysis, TP53 mutations were classified as G>T transversion hotspots or CpG transition hotspots, following the established association of the former with tobacco exposure and the latter with spontaneous deamination (S8,S9) and the hotspot frequencies catalogued in the IARC TP53 database and in systematic functional analyses of missense p53 variants (S10,S11); the alleles assigned to each class are listed in the Supplementary Methods. Amplification was defined as GISTIC +2 and deep deletion as GISTIC-2 (S6). For the cross-cohort prevalence analysis, genes were additionally classified by how a variant call is made rather than by pathway: oncogene calls are hotspot lookups requiring no interpretation, whereas tumor suppressor calls require deciding whether a novel truncating or missense variant is pathogenic.

### Choice of EGFR, ALK, and ROS1 as the driver-positive definition

Regulatory approvals for the seven targetable drivers not used to define driver-positive disease fell during or after the November 2017 to December 2023 accrual window: larotrectinib in 2018, entrectinib in 2019, capmatinib and selpercatinib in 2020, sotorasib in 2021, adagrasib and trastuzumab deruxtecan in 2022, and zenocutuzumab in 2024. Access to matched therapy for those alterations was therefore neither uniform across the cohort nor present for its earlier years, whereas first-line tyrosine kinase inhibitors for EGFR-, ALK-, and ROS1-altered disease were guideline-endorsed standard of care throughout. Each of the remaining drivers is also present in fewer than 5% of tumors, too few to support stratified survival modeling.

### Drivers retained within the driver-negative group

RET fusions, HER2 activating mutations, NTRK fusions, and NRG1 fusions are the alterations biologically closest to EGFR, ALK, and ROS1: each acts through a receptor tyrosine kinase, each defines an oncogene-addicted tumor typically dependent on a single signaling axis, and each is enriched among never-smokers with adenocarcinoma, so their presence in the comparator group biases the driver-positive versus driver-negative contrast toward the null. Because StrataNGS reports RNA-based fusions these alterations are ascertainable in the discovery cohort, and they were uncommon. KRAS G12C, MET exon 14 skipping, and BRAF V600E tumors occur predominantly in patients with a smoking history and are associated with higher tumor mutational burden; at the gene level, KRAS, MET, and BRAF pathogenic variants were present in 820 (36.6%), 149 (6.6%), and 133 (5.9%) of driver-negative patients respectively, and the G12C, exon 14 skipping, and V600E subsets are smaller than these gene-level counts.

### Validation cohort assembly

The non-redundant cohort of 209,672 unique samples was constructed by normalizing sample identifiers, assigning each sample to a single contributing study, and excluding cell lines, xenografts, and liquid biopsies. The archives include the disease-focused sequencing consortia and the registry-scale aggregations of clinical panel data (1,2,S4). Because patient-level demographics are not carried in the aggregated clinical extract, we re-extracted the clinical patient and sample files of 50 lung-contributing archives, recovering age, sex, race, ethnicity, smoking history, stage, and sample type where each study recorded them, and harmonized the differing vocabularies used across studies.

### Allele and fusion definitions in the validation cohort

EGFR activating alleles were defined as exon 19 in-frame deletions spanning codons 729-761, L858R, L861Q/R, G719A/C/S/D, S768I, and exon 20 in-frame insertions spanning codons 762-775. Oncogene protein-level and fusion records were re-extracted from all 37 lung-contributing archives carrying mutation data, yielding 39,374 oncogene mutation records and 4,127 fusion records, with intragenic ALK events excluded. RET, NTRK1/2/3, and NRG1 fusions were called from the structural-variant files of the 18 contributing studies that deposit them, requiring two distinct partner genes and excluding intragenic, antisense, intergenic, and within-transcript rearrangements; 9,052 of the 10,788 adenocarcinomas came from those studies. ERBB2 activating alleles were defined as exon 20 in-frame insertions and duplications spanning codons 770 to 783, in-frame indels at the exon 19 L755-N758 hotspot, and the recurrent transmembrane and kinase-domain missense alleles S310F/Y, L755S/P/A, D769H/Y, G776V/C and G776delins, V777L/M, V842I, L869R, V659E/D, G660D, and R896C; R678Q, I767M, and singleton missense changes of uncertain significance were counted only in the gene-level call. An EGFR variant was assigned to the exon 20 insertion class if it was an in-frame insertion with a leading codon between 762 and 775, and to the non-exon-20 activating class if it was L858R, L861Q/R, S768I, or G719A/C/S/D, an in-frame indel with a leading codon between 729 and 761, or one of the delins forms encoding a classical allele; the two classes are mutually exclusive and together reproduce the EGFR-activating group used elsewhere.

### Reconciliation of tumor counts and survival conventions

The three mutually exclusive adenocarcinoma groups comprise a base of 10,788 tumors from 31 studies. The nine tumors separating this from the 10,797 used in the prevalence analyses come from three studies whose mutation files could not be re-parsed at protein level, and a further four tumors carrying both an EGFR-activating allele and an ALK fusion are excluded from the mutually exclusive grouping, so the three groups sum to 10,784. Survival analyses of the driver groups were performed one row per patient rather than one row per patient and study, because several contributing releases from the same institution are nested and would otherwise count the same patient, and the same death, more than once.

### TP53 hotspot classification

G>T transversion hotspots were defined as V157F, R158L/P, and R273L, and CpG transition hotspots as R175H, R248W/Q, R273H/C, R282W, and R213*.

### Exon 20 insertion classes in the discovery cohort

The discovery exon 20 insertion classes were drawn to match the validation definitions: an ERBB2 or EGFR variant was assigned to the insertion class only if it was an in-frame insertion within the codon window used for that gene in the validation cohort. Non-insertion exon 20 variants, including EGFR T790M, are assigned to the non-exon-20 class. Under this definition the EGFR insertion class comprises 5.7% of EGFR-mutant patients, against 6.2% of the EGFR-activating group in the validation cohort; a broader definition admitting any exon 20 variant assigns 14.8%. TP53 status was compared between classes by chi-square test, and survival within each class by Cox regression with delayed entry at the date of NGS and TP53 as the only term. Comparisons between discovery and validation estimates are informal: the adjustment sets differ, the validation models are stratified by contributing study, and the exposure is a clinically reported TP53 pathogenic variant in discovery against TP53 alteration including deep deletion in validation.

## Supplementary Tables

**Supplementary Table S1.** Comparison of the discovery and validation cohorts. The two cohorts differ in almost every respect except the disease. The discovery cohort holds stage and treatment status constant and records comorbidity and performance status but not smoking; the validation cohort records smoking, stage, and sample type but neither comorbidity, performance status, nor treatment.

| Domain | Discovery (institutional) | Validation (public) |
| --- | --- | --- |
| Design | one integrated healthcare organization | 483 aggregated study archives |
| Contributing studies | 1 | 37 |
| Adenocarcinoma | 2,797 | 10,797 |
| Squamous | 498 | 2,185 |
| Stage | stage IV throughout (100%) | recorded for 49.3%; 30.0% stage IV |
| First-line systemic therapy | 97% treated; regimen class recorded | not recorded |
| Assay | one targeted panel | 21.1% whole exome, rest panel |
| Age recorded | 100% | 90.4% |
| Sex recorded | 100% | 96.7% |
| Race recorded | 95.1% | 61.7% |
| Comorbidity | 100% | not recorded |
| Performance status | 9.0% | not recorded |
| Smoking | not recorded | 32.3% |
| Sample type | not extracted | 80.8% (35.4% metastasis) |
| Median age, adenocarcinoma | 69 | 66.2 |
| Female, adenocarcinoma | 58.6% | 58.5% |
| Asian, adenocarcinoma | 27.8% | 12.4% |
| Deaths | 2,291 | 2,828 (adenocarcinoma) |
| Survival model | Cox, delayed entry, 33-36 covariates | Cox, stratified by study |
| Variant calling | clinical pathogenic-variant report | any protein-altering variant |
| Driver definition | gene-level EGFR/ALK/ROS1 | EGFR alleles; ALK fusion |

**Supplementary Table S2.** Replication of the differential association. The ratio is the TP53 hazard ratio in driver-positive divided by that in driver-negative disease; a ratio of 1 would indicate that TP53 carries equal prognostic weight with and without a targetable driver. Cohorts were pooled by fixed-effect inverse-variance weighting and heterogeneity was tested by a two-sided z test on the difference of log ratios.

| Cohort | N | Ratio of TP53 hazard ratios [95% CI] | p |
| --- | --- | --- | --- |
| Discovery | 3,295 | 1.66 [1.34-2.05] | <0.001 |
| Validation | 7,571 | 1.74 [1.44-2.10] | <0.001 |
| Fixed-effect pooled | 10,866 | 1.70 [1.48-1.96] | <0.001 |
| Heterogeneity between cohorts |  |  | 0.74 |

**Supplementary Table S3.** TP53 by driver interaction across adjustment models in the validation cohort. All models are Cox regression models stratified by contributing study with a multiplicative interaction term. Rows three and four comprise identical patients and differ only in whether a never-smoker term is included.

| Model | n | deaths | TP53 by driver interaction [95% CI] | p |
| --- | --- | --- | --- | --- |
| Unadjusted | 7,558 | 2,827 | 1.73 [1.45-2.07] | <0.001 |
| + age, sex | 7,333 | 2,753 | 1.77 [1.47-2.11] | <0.001 |
| Smoking subset, no smoking term | 3,120 | 1,005 | 1.661 [1.228-2.247] | 0.001 |
| Same subset, + smoking | 3,120 | 1,005 | 1.658 [1.226-2.242] | 0.001 |
| + smoking, age, sex | 3,067 | 992 | 1.72 [1.27-2.32] | <0.001 |
| Stage subset, + stage | 3,332 | 1,087 | 1.57 [1.18-2.08] | 0.002 |

**Supplementary Table S4.** Sixteen survival associations estimated separately in each cohort. Discovery estimates are fully adjusted; validation estimates are mutually adjusted and stratified by contributing study. Pooled estimates are fixed-effect inverse-variance weighted and heterogeneity was tested by a two-sided z test on the difference of log hazard ratios.

| Association | Discovery HR [95% CI] | Validation HR [95% CI] | Pooled | p het. |
| --- | --- | --- | --- | --- |
| TP53 altered (adenocarcinoma) | 1.30 [1.18-1.44] | 1.53 [1.41-1.66] | 1.43 [1.35-1.53] | 0.013 |
| Driver positive | 0.73 [0.64-0.83] | 0.79 [0.72-0.88] | 0.77 [0.71-0.83] | 0.31 |
| KRAS mutated | 1.05 [0.93-1.19] | 1.10 [1.00-1.20] | 1.08 [1.00-1.16] | 0.57 |
| STK11 mutated | 1.24 [0.99-1.56] | 1.31 [1.17-1.46] | 1.30 [1.17-1.43] | 0.68 |
| KEAP1 mutated | 1.31 [1.01-1.72] | 1.46 [1.30-1.63] | 1.44 [1.30-1.59] | 0.48 |
| SMARCA4 mutated | 1.38 [0.99-1.91] | 1.42 [1.25-1.61] | 1.41 [1.25-1.59] | 0.87 |
| MYC amplified | 1.45 [1.17-1.79] | 1.47 [1.27-1.70] | 1.46 [1.29-1.65] | 0.93 |
| MDM2 amplified | 1.10 [0.86-1.42] | 1.17 [0.99-1.38] | 1.15 [1.00-1.32] | 0.70 |
| RB1 altered | 1.14 [0.93-1.39] | 1.15 [0.99-1.35] | 1.15 [1.01-1.30] | 0.92 |
| NF1 mutated | 0.98 [0.73-1.33] | 0.84 [0.72-0.97] | 0.87 [0.76-0.99] | 0.36 |
| BRAF mutated | 1.00 [0.81-1.24] | 0.99 [0.84-1.16] | 0.99 [0.87-1.13] | 0.94 |
| PIK3CA mutated | 1.14 [0.92-1.41] | 0.98 [0.84-1.16] | 1.04 [0.91-1.18] | 0.30 |
| TP53 altered (squamous) | 0.97 [0.74-1.28] | 0.93 [0.74-1.17] | 0.95 [0.80-1.13] | 0.81 |
| KEAP1 mutated (squamous) | 0.70 [0.36-1.36] | 1.25 [0.99-1.60] | 1.17 [0.93-1.47] | 0.10 |
| STK11 mutated (squamous) | 6.14 [2.29-16.47] | 1.71 [1.02-2.86] | 2.25 [1.42-3.54] | 0.024 |
| CDKN2A altered | 1.32 [1.18-1.48] | 1.74 [1.55-1.95] | 1.52 [1.41-1.65] | <0.001 |

**Supplementary Table S5.**
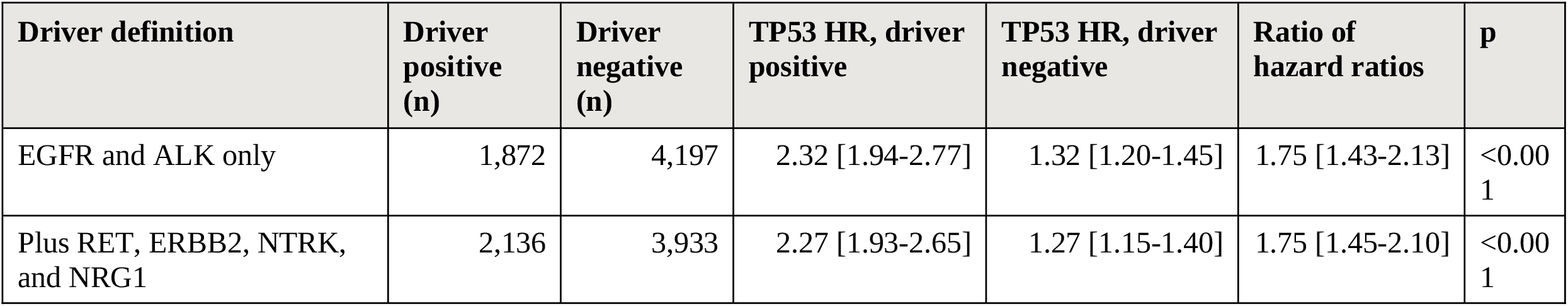
Sensitivity of the differential association to the definition of a targetable driver in the cBioPortal validation cohort. Both rows are estimated in the same 6,069 cBioPortal validation adenocarcinoma patients with survival data; only the assignment of patients to the driver-positive group changes. The ratio is estimated as a multiplicative interaction term in a Cox regression model stratified by contributing study.

**Supplementary Table S6.** Sensitivity of the TP53 by EGFR exon 20 insertion interaction. All models are Cox regression models stratified by contributing study, fitted to the EGFR exon 20 insertion group and the no-targetable-driver group only, with a multiplicative interaction term. Restricting the reference to studies that deposit structural variants removes tumors that cannot be shown to be free of a fusion driver. Smoking status is imputed rather than observed for most of these patients.

| Model | n | EGFR exon 20 insertion (n) | Ratio of hazard ratios | p |
| --- | --- | --- | --- | --- |
| Primary | 4,606 | 141 | 3.88 [1.96-7.71] | <0.001 |
| Reference restricted to structural-variant studies | 4,031 | 134 | 3.39 [1.69-6.81] | 0.001 |
| Smoking subset, no smoking term | 4,323 | 132 | 4.00 [1.95-8.22] | <0.001 |
| Same subset, plus ever-smoker term | 4,323 | 132 | 3.97 [1.93-8.15] | <0.001 |
| Never smokers only | 666 | 78 | 3.04 [1.07-8.67] | 0.037 |

**Supplementary Table S7.** Characteristics of the discovery cohort by histology. Values are n (%) unless otherwise stated. PV, pathogenic variant; WT, wild-type. Gene and TP53 percentages are of the histology column; race percentages are of patients in whom race is recorded (n = 2,651 adenocarcinoma and n = 482 squamous). p values are from chi-square tests for categorical variables and Wilcoxon rank-sum tests for continuous variables. The full gene list is provided as supplementary data file D7.

| Characteristic |  | Adenocarcinoma (n = 2,797) | Squamous (n = 498) | p |
| --- | --- | --- | --- | --- |
| Age (years) | median | 69.0 | 73.0 | <0.001 |
|  | range | 23-96 | 43-93 |  |
| Race | White | 1,351 (51.0%) | 313 (64.9%) | <0.001 |
|  | Asian | 736 (27.8%) | 70 (14.5%) | <0.001 |
|  | Black | 222 (8.4%) | 55 (11.4%) | <0.001 |
|  | Hispanic | 113 (4.3%) | 17 (3.5%) | <0.001 |
|  | Other | 229 (8.6%) | 27 (5.6%) | <0.001 |
| Sex | female | 1,640 (58.6%) | 213 (42.8%) | <0.001 |
| Charlson index | median | 1.0 | 2.0 | <0.001 |
| Performance status | ECOG 0-1 | 218 (7.8%) | 37 (7.4%) | 0.242 |
|  | ECOG 2-4 | 31 (1.1%) | 10 (2.0%) | 0.242 |
|  | unknown | 2,548 (91.1%) | 451 (90.6%) | 0.242 |
| TP53 | WT | 1,361 (48.7%) | 106 (21.3%) | <0.001 |
|  | PV | 1,436 (51.3%) | 392 (78.7%) | <0.001 |
| EGFR | PV | 852 (30.5%) | 35 (7.0%) | <0.001 |
| KRAS | PV | 805 (28.8%) | 36 (7.2%) | <0.001 |
| CDKN2A | PV | 576 (20.6%) | 179 (35.9%) | <0.001 |
| PIK3CA | PV | 136 (4.9%) | 117 (23.5%) | <0.001 |
| ALK | PV | 131 (4.7%) | 2 (0.4%) | <0.001 |
| STK11 | PV | 128 (4.6%) | 5 (1.0%) | <0.001 |
| MYC | amplification | 123 (4.4%) | 18 (3.6%) | 0.426 |
| KEAP1 | PV | 86 (3.1%) | 17 (3.4%) | 0.689 |
| SMARCA4 | PV | 53 (1.9%) | 7 (1.4%) | 0.452 |
| ROS1 | PV | 34 (1.2%) | 0 (0.0%) | 0.013 |

**Supplementary Table S8.** Characteristics of the discovery cohort by targetable driver status. Values are n (%) unless otherwise stated. Driver-positive denotes a pathogenic EGFR, ALK, or ROS1 alteration. The groups differ on almost every demographic and genomic characteristic but not on TP53 status, MYC amplification, or RB1. p values are from chi-square tests for categorical variables and Wilcoxon rank-sum tests for continuous variables.

| Characteristic |  | EGFR/ALK/ROS1 positive (n = 1,053) | negative (n = 2,242) | p |
| --- | --- | --- | --- | --- |
| Age (years) | median | 67.0 | 71.0 | <0.001 |
| Race | White | 358 (35.6%) | 1,306 (61.4%) | <0.001 |
|  | Asian | 457 (45.5%) | 349 (16.4%) | <0.001 |
|  | Black | 42 (4.2%) | 235 (11.0%) | <0.001 |
| Sex | female | 690 (65.5%) | 1,163 (51.9%) | <0.001 |
| Charlson index | median | 1.0 | 2.0 | <0.001 |
| TP53 | WT | 492 (46.7%) | 975 (43.5%) | 0.084 |
|  | PV | 561 (53.3%) | 1,267 (56.5%) | 0.084 |
| KRAS | PV | 21 (2.0%) | 820 (36.6%) | <0.001 |
| STK11 | PV | 0 (0.0%) | 133 (5.9%) | <0.001 |
| CDKN2A | PV | 268 (25.5%) | 487 (21.7%) | 0.017 |
| MDM2 | amplification | 82 (7.8%) | 75 (3.3%) | <0.001 |
| MYC | amplification | 47 (4.5%) | 94 (4.2%) | 0.720 |
| RB1 | PV | 67 (6.4%) | 126 (5.6%) | 0.397 |
| BRAF | PV | 12 (1.1%) | 133 (5.9%) | <0.001 |
| CTNNB1 | PV | 77 (7.3%) | 45 (2.0%) | <0.001 |

**Supplementary Table S9.** Hazard ratios for OS associated with TP53 pathogenic variants, by targetable driver status and histology. WT, wild-type. n denotes the patients contributing to each survival model rather than the tumors in the group. Discovery estimates are adjusted for age, race, sex, comorbidity, performance status, histology, and 25 additional genes. Validation estimates are given both stratified by contributing study alone and mutually adjusted for 15 to 17 additional genes.

| Cohort | Group | Model | n | TP53 HR [95% CI] | p |
| --- | --- | --- | --- | --- | --- |
| Discovery | EGFR/ALK/ROS1 positive | adjusted | 1,053 | 1.80 [1.50-2.15] | <0.001 |
| Discovery | EGFR/ALK/ROS1 negative | adjusted | 2,242 | 1.08 [0.97-1.21] | 0.15 |
| Discovery | adenocarcinoma (all) | adjusted | 2,797 | 1.30 [1.18-1.44] | <0.001 |
| Discovery | squamous | adjusted | 498 | 0.97 [0.74-1.28] | 0.85 |
| Validation | EGFR-activating | study-stratified | 2,061 | 2.27 [1.92-2.69] | <0.001 |
| Validation | ALK fusion | study-stratified | 240 | 2.42 [1.42-4.10] | 0.001 |
| Validation | EGFR/ALK WT | study-stratified | 5,268 | 1.30 [1.20-1.42] | <0.001 |
| Validation | adenocarcinoma (all) | study-stratified | 7,571 | 1.48 [1.37-1.59] | <0.001 |
| Validation | adenocarcinoma (all) | mutually adjusted | 7,314 | 1.53 [1.41-1.66] | <0.001 |
| Validation | squamous | study-stratified | 1,446 | 0.97 [0.78-1.20] | 0.78 |
| Validation | squamous | mutually adjusted | 1,407 | 0.93 [0.74-1.17] | 0.54 |

**Supplementary Table S10.** TP53 alteration and overall survival by targetable driver group in cBioPortal validation adenocarcinoma. Prevalence is among the 9,052 adenocarcinomas from studies that deposited structural variants; odds ratios are Fisher exact against the no-targetable-driver group with Benjamini-Hochberg control across the six comparisons. Survival estimates are from Cox regression models stratified by contributing study in 6,069 patients with survival data, analyzed one row per patient. The ratio is the TP53 hazard ratio within the group divided by that in the no-targetable-driver group, estimated as a multiplicative interaction term in a model fitted to that group and the no-driver group only. Groups are mutually exclusive; 29 tumors carrying more than one driver are excluded from the group rows. Estimates are reported as not estimable where fewer than five deaths occurred among TP53-altered patients.

| Driver group | n | TP53 altered (%) | OR vs no driver | Driver HR vs no driver | TP53 HR within group | Ratio vs no driver |
| --- | --- | --- | --- | --- | --- | --- |
| EGFR-activating | 2,900 | 53.1 | 1.39 [1.27-1.52] | 0.67 [0.61-0.74] | 2.32 [1.91-2.81] | 1.81 [1.46-2.24] |
| ALK fusion | 360 | 26.4 | 0.44 [0.35-0.56] | 0.56 [0.43-0.73] | 2.59 [1.47-4.54] | 1.93 [1.12-3.33] |
| RET fusion | 145 | 28.3 | 0.48 [0.34-0.70] | 0.70 [0.50-0.99] | 3.09 [1.47-6.47] | 2.78 [1.39-5.55] |
| ERBB2 activating | 248 | 48.8 | 1.17 [0.91-1.51] | 1.20 [0.95-1.52] | 1.96 [1.21-3.15] | 1.34 [0.83-2.14] |
| NTRK fusion | 16 | 18.8 | 0.28 [0.08-0.99] | not estimable | not estimable | not estimable |
| NRG1 fusion | 18 | 22.2 | 0.35 [0.12-1.07] | 0.78 [0.32-1.90] | not estimable | not estimable |
| No targetable driver | 5,336 | 44.9 | reference | reference | 1.27 [1.15-1.40] | reference |

**Supplementary Table S11.**
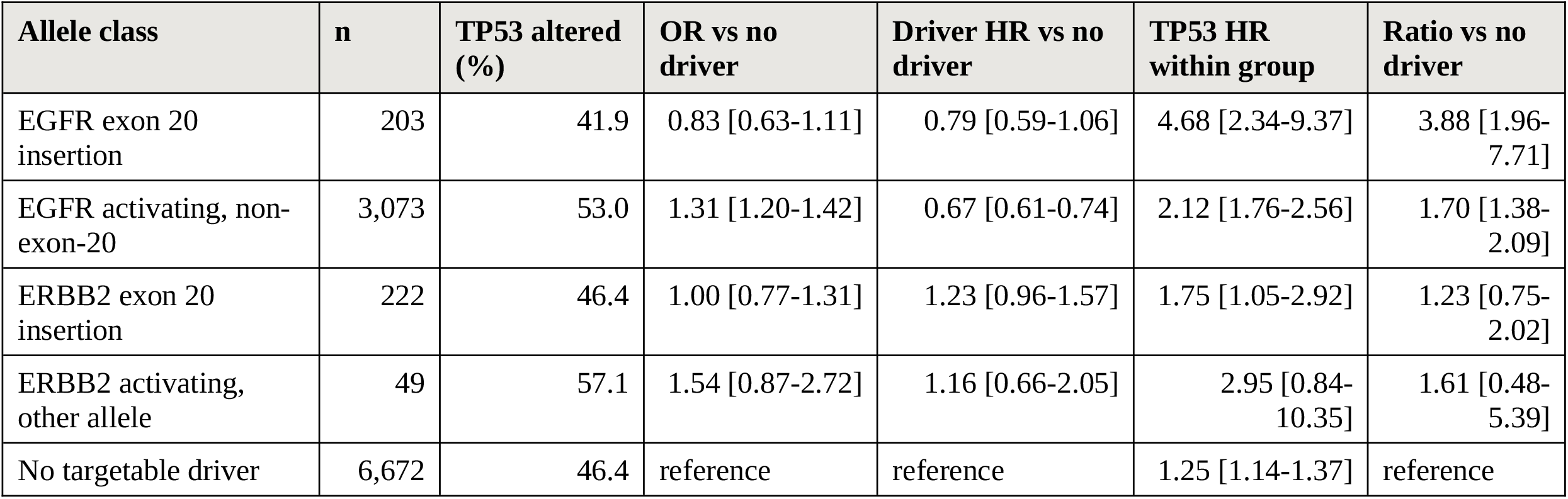
TP53 alteration and overall survival by exon 20 insertion status in cBioPortal validation adenocarcinoma. Prevalence is among 10,788 adenocarcinomas; odds ratios are Fisher exact against the no-targetable-driver group with Benjamini-Hochberg control. Survival estimates are from Cox regression models stratified by contributing study in 6,798 patients with survival data, analyzed one row per patient. The ratio is the TP53 hazard ratio within the group divided by that in the no-targetable-driver group, estimated as a multiplicative interaction term fitted to that group and the no-driver group only. Groups are mutually exclusive; 30 tumors carrying more than one driver are excluded. The ERBB2 other-allele row rests on 25 patients with 12 deaths and should be read with that in mind.

| Allele class | n | TP53 altered (%) | OR vs no driver | Driver HR vs no driver | TP53 HR within group | Ratio vs no driver |
| --- | --- | --- | --- | --- | --- | --- |
| EGFR exon 20 insertion | 203 | 41.9 | 0.83 [0.63-1.11] | 0.79 [0.59-1.06] | 4.68 [2.34-9.37] | 3.88 [1.96-7.71] |
| EGFR activating, non-exon-20 | 3,073 | 53.0 | 1.31 [1.20-1.42] | 0.67 [0.61-0.74] | 2.12 [1.76-2.56] | 1.70 [1.38-2.09] |
| ERBB2 exon 20 insertion | 222 | 46.4 | 1.00 [0.77-1.31] | 1.23 [0.96-1.57] | 1.75 [1.05-2.92] | 1.23 [0.75-2.02] |
| ERBB2 activating, other allele | 49 | 57.1 | 1.54 [0.87-2.72] | 1.16 [0.66-2.05] | 2.95 [0.84-10.35] | 1.61 [0.48-5.39] |
| No targetable driver | 6,672 | 46.4 | reference | reference | 1.25 [1.14-1.37] | reference |

**Supplementary Table S12.** TP53 pathogenic variants and overall survival within the exon 20 insertion classes of the KPNC discovery cohort. Percentages are within the insertion class, which comprises in-frame insertions only and excludes EGFR T790M. Medians are Kaplan-Meier estimates with 95% confidence intervals, measured from the diagnosis of advanced disease with delayed entry at the date of NGS. Hazard ratios are from Cox regression with delayed entry and TP53 as the only term and are therefore unadjusted; the p value is the Wald test on that term. TP53 prevalence in the corresponding non-exon-20 classes was 73.5% for ERBB2 (n = 49) and 59.0% for EGFR (n = 836), giving p = 0.020 and p = 0.051 against the insertion classes by chi-square test.

| Insertion class | n | TP53 PV n (%) | Deaths | Median OS, TP53 WT (months) | Median OS, TP53 PV (months) | TP53 HR [95% CI] |
| --- | --- | --- | --- | --- | --- | --- |
| ERBB2 exon 20 insertion | 60 | 31 (51.7) | 47 | 19.2 [12.7-33.0] | 14.0 [8.1-48.7] | 0.87 [0.48-1.57], p = 0.65 |
| EGFR exon 20 insertion | 51 | 23 (45.1) | 43 | 19.3 [12.9-35.5] | 18.1 [11.6-45.1] | 1.25 [0.68-2.30], p = 0.47 |

**Supplementary Figure S1.**
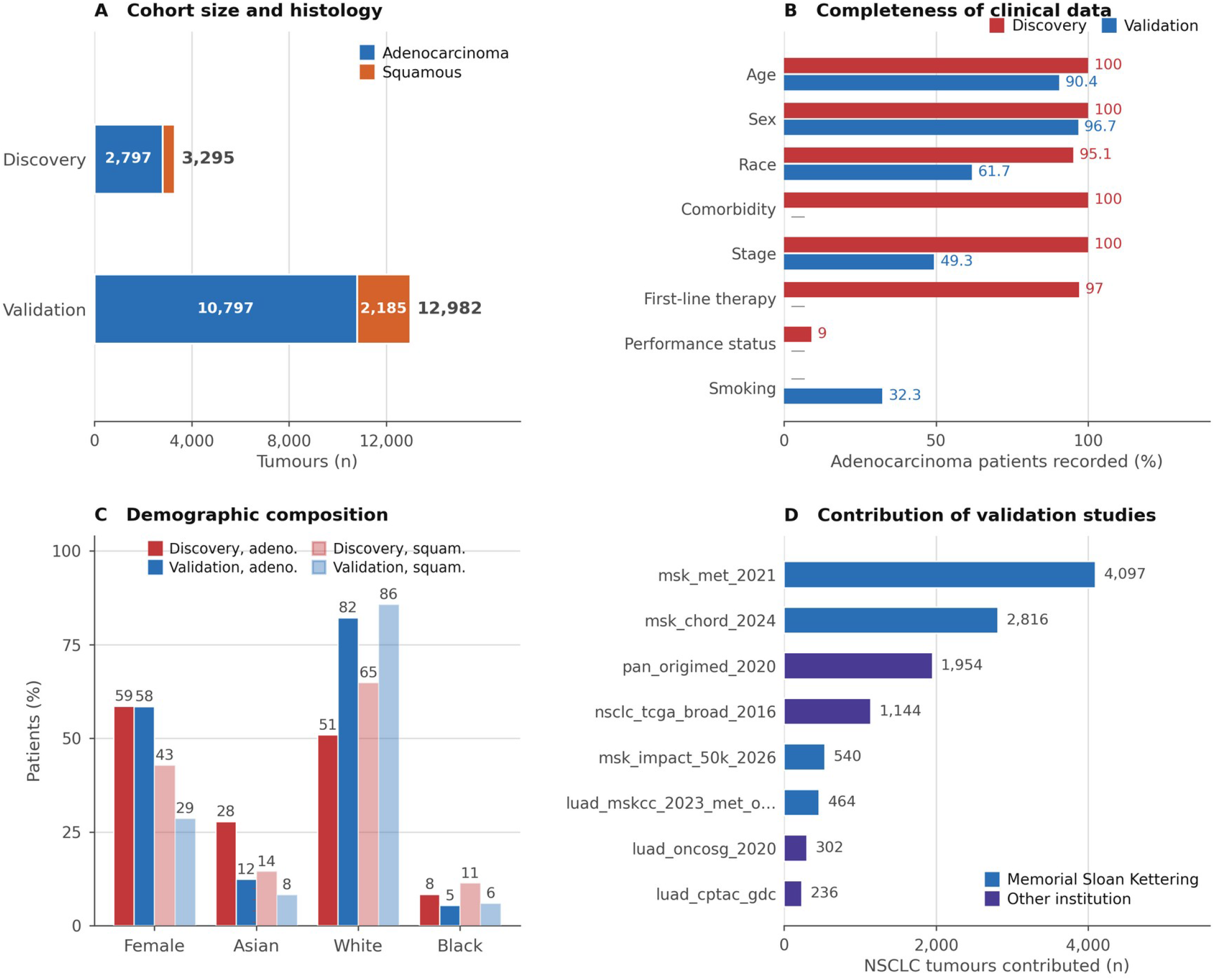
Composition and data completeness of the two cohorts. (A) Number of tumors by histology. (B) Percentage of patients with adenocarcinoma for whom each variable is recorded; the discovery cohort records comorbidity, performance status, stage, and treatment but not smoking, whereas the validation cohort records smoking, stage, and sample type but none of the others. (C) Demographic composition by cohort and histology, among patients in whom the variable is recorded; the adenocarcinoma sex distribution is nearly identical between cohorts (58.6% versus 58.5% female) whereas race is not (27.8% versus 12.4% Asian). (D) The eight largest contributing validation studies; 37 studies contribute in total, the ten largest supply 92% of the tumors, and studies from a single institution supply 63%, which is why every validation survival model is stratified by contributing study. Discovery cohort n = 3,295; validation cohort n = 12,982 from 37 studies. Small cell carcinoma is excluded from both cohorts.

**Supplementary Figure S2.**
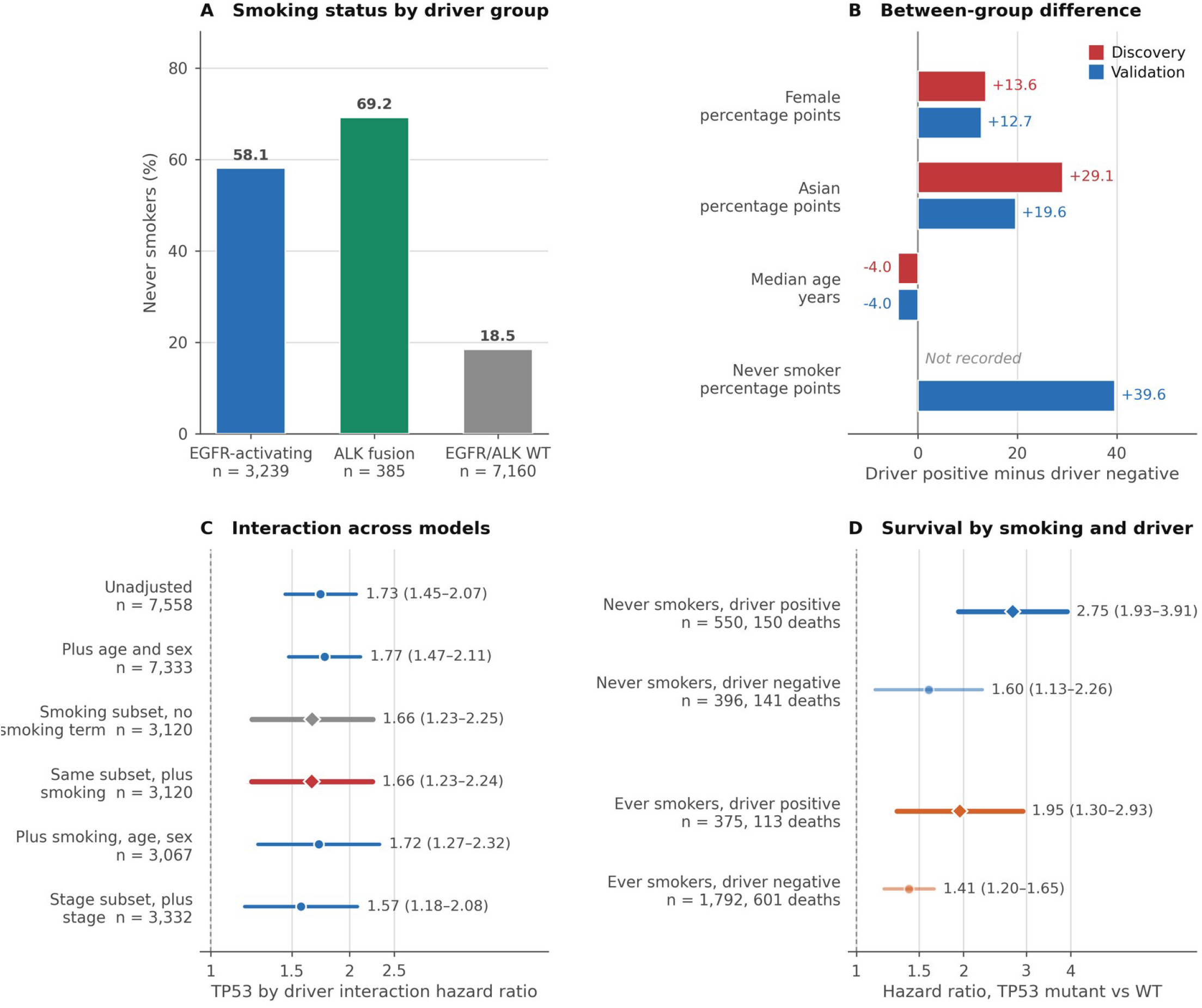
Smoking history and the TP53 PV by driver interaction in the cBioPortal validation cohort. (A) Proportion of never smokers by driver group; smoking status was recorded for 28%, 20%, and 35% of the EGFR-activating, ALK-fusion, and EGFR/ALK wild-type groups respectively, and against wild-type disease the odds ratios for never smoking were 6.12 and 9.93 (both p < 0.001). (B) Difference between driver-positive and driver-negative groups for each characteristic, in percentage points or in years, in each cohort. (C) TP53 by driver interaction hazard ratio across successive adjustment models; the two highlighted rows comprise identical patients and differ only in whether a never-smoker term is included, and the estimate moves from 1.661 to 1.658. (D) Hazard ratio for TP53 pathogenic variants within each combination of smoking status and driver status. All models are Cox regression models stratified by contributing study, with a multiplicative interaction term. Horizontal bars are 95% confidence intervals. Smoking history was recovered for 32.3% of patients with adenocarcinoma and for 3,120 of those with survival data.

## Supplementary Data Files

D1. TP53 prevalence by histology and by driver group, validation cohort.

D2. TP53 mutation spectrum by histology and by driver group.

D3. Co-alteration with TP53, burden-stratified, by histology and driver group.

D4. Driver prevalence by histology and driver group definitions.

D5. TP53 survival estimates by histology and driver group.

D6. Mutually adjusted survival models by histology and driver group.

D7. Discovery cohort characteristics by histology and by driver status.

D8. Validation cohort demographics by histology and by driver group, with tests.

D9. Validation cohort study composition.

D10. Prevalence concordance, 26 comparisons, and the variant call class summary.

D11. Survival association concordance, 16 comparisons, with pooled estimates.

D12. TP53 by driver status and the interaction replication.

D13. Mixture estimate for the discovery squamous label.

D14. Smoking-adjusted interaction models and stratum-specific estimates.

D15. Cohort side-by-side summary.

D16. RET, ERBB2, NTRK, and NRG1 prevalence, TP53 co-alteration, demographics, and survival.

D17. ERBB2 and EGFR exon 20 insertion allele spectrum, TP53 co-alteration, and survival.

## Supplementary Data Files

**Supplementary Data File D1.**
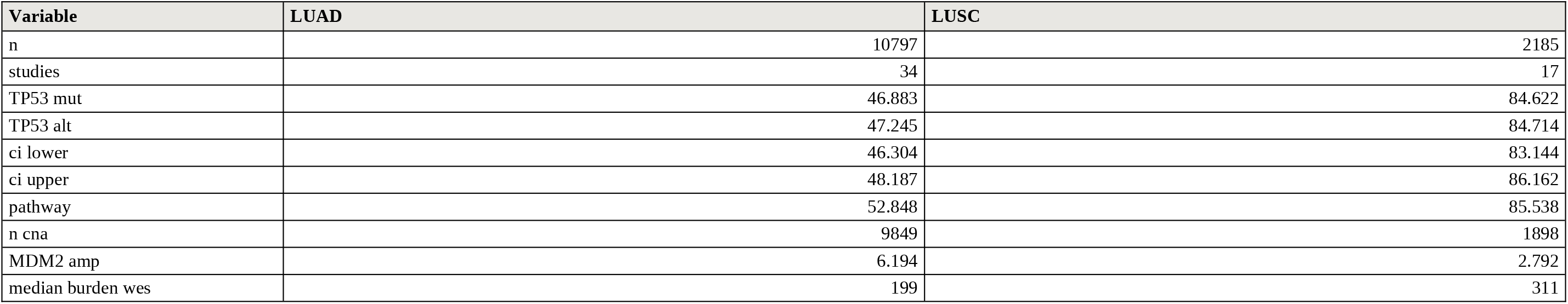
TP53 prevalence by histology, validation cohort.

**Supplementary Data File D1 (continued).** TP53 alteration by driver group within validation adenocarcinoma.

| Variable | EGFR-activating | ALK fusion | EGFR/ALK wild-type |
| --- | --- | --- | --- |
| n | 3239 | 385 | 7160 |
| TP53 alt | 52.454 | 25.714 | 46.047 |
| ci lower | 50.733 | 21.603 | 44.895 |
| ci upper | 54.17 | 30.305 | 47.204 |
| n altered | 1699 | 99 | 3297 |
| median burden wes | 94 | 1,798.5 | 279 |

**Supplementary Data File D2.**
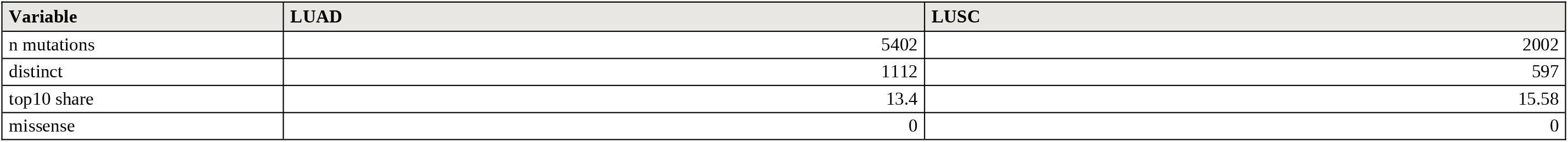
TP53 mutation spectrum by histology, validation cohort.

| Variable | LUAD | LUSC |
| --- | --- | --- |
| n mutations | 5402 | 2002 |
| distinct | 1112 | 597 |
| top10 share | 13.4 | 15.58 |
| missense | 0 | 0 |

**Supplementary Data File D2 (continued).** TP53 mutation spectrum by driver group.

| Variable | EGFR-activating | ALK fusion | EGFR/ALK wild-type |
| --- | --- | --- | --- |
| n mutations | 1775 | 101 | 3519 |
| distinct | 615 | 77 | 830 |
| transversion hotspots | 2.535 | 0 | 6.905 |
| CpG hotspots | 10.93 | 16.832 | 6.621 |
| top | R273C 2.4%, R273H 2.1%, R248W 1.5%, R213* 1.4% | R175H 5.9%, Q331* 5.0%, R248W 4.0%, R248Q 3.0% | V157F 2.3%, R158L 2.3%, R273L 2.0%, G154V 1.2% |

**Supplementary Data File D3.** Co-alteration with TP53 by histology, burden-stratified Cochran-Mantel-Haenszel odds ratios.

| histology | gene | n | n alt | % | % in TP53alt | % in TP53wt | CMH OR | p | q |
| --- | --- | --- | --- | --- | --- | --- | --- | --- | --- |
| LUAD | KRAS | 10767 | 2990 | 27.77 | 21.409 | 33.457 | 0.388 | <0.001 | <0.001 |
| LUAD | EGFR | 10764 | 3528 | 32.776 | 36.907 | 29.079 | 1.703 | <0.001 | <0.001 |
| LUAD | STK11 | 10775 | 1429 | 13.262 | 9.957 | 16.224 | 0.386 | <0.001 | <0.001 |
| LUAD | KEAP1 | 10584 | 1235 | 11.669 | 11.313 | 11.989 | 0.64 | <0.001 | <0.001 |
| LUAD | SMARCA4 | 10584 | 766 | 7.237 | 8.42 | 6.174 | 1.002 | 0.986 | 0.986 |
| LUAD | RB1 | 10774 | 515 | 4.78 | 8.563 | 1.39 | 5.408 | <0.001 | <0.001 |
| LUAD | NF1 | 10736 | 773 | 7.2 | 10.344 | 4.392 | 1.824 | <0.001 | <0.001 |
| LUAD | BRAF | 10729 | 556 | 5.182 | 5.55 | 4.854 | 0.925 | 0.416 | 0.485 |
| LUAD | PIK3CA | 10776 | 571 | 5.299 | 6.031 | 4.643 | 1.077 | 0.431 | 0.485 |
| LUAD | MET | 10715 | 400 | 3.733 | 3.878 | 3.604 | 0.975 | 0.86 | 0.894 |
| LUAD | ERBB2 | 10682 | 418 | 3.913 | 4.378 | 3.497 | 1.244 | 0.041 | 0.058 |
| LUAD | ATM | 10744 | 790 | 7.353 | 5.319 | 9.174 | 0.378 | <0.001 | <0.001 |
| LUAD | ARID1A | 10609 | 613 | 5.778 | 7.677 | 4.067 | 1.451 | <0.001 | <0.001 |
| LUAD | NFE2L2 | 10452 | 148 | 1.416 | 1.699 | 1.162 | 1.085 | 0.696 | 0.751 |
| LUAD | PTEN | 10694 | 256 | 2.394 | 3.106 | 1.756 | 1.458 | 0.005 | 0.008 |
| LUAD | KMT2D | 10505 | 596 | 5.674 | 7.535 | 3.998 | 1.425 | <0.001 | <0.001 |
| LUAD | CDKN2A | 10747 | 511 | 4.755 | 7.092 | 2.663 | 2.246 | <0.001 | <0.001 |
| LUAD | APC | 10779 | 526 | 4.88 | 6.007 | 3.87 | 1.187 | 0.073 | 0.099 |
| LUAD | NOTCH1 | 10630 | 313 | 2.945 | 4.266 | 1.764 | 1.791 | <0.001 | <0.001 |
| LUAD | CREBBP | 10524 | 376 | 3.573 | 4.332 | 2.889 | 1.127 | 0.296 | 0.363 |
| LUAD | EP300 | 10502 | 246 | 2.342 | 3.199 | 1.573 | 1.521 | 0.003 | 0.005 |
| LUAD | PTPRD | 9120 | 821 | 9.002 | 12.488 | 5.903 | 1.625 | <0.001 | <0.001 |
| LUAD | SETD2 | 10547 | 573 | 5.433 | 5.142 | 5.695 | 0.717 | <0.001 | <0.001 |
| LUAD | CTNNB1 | 10713 | 386 | 3.603 | 3.008 | 4.135 | 0.581 | <0.001 | <0.001 |
| LUAD | MGA | 8912 | 487 | 5.465 | 6.622 | 4.434 | 1.137 | 0.205 | 0.263 |
| LUAD | U2AF1 | 10503 | 226 | 2.152 | 1.889 | 2.388 | 0.628 | 0.001 | 0.002 |
| LUAD | CIC | 10538 | 193 | 1.832 | 2.602 | 1.137 | 1.695 | <0.001 | 0.002 |
| LUSC | KRAS | 2185 | 85 | 3.89 | 3.133 | 8.084 | 0.304 | <0.001 | <0.001 |
| LUSC | EGFR | 2185 | 92 | 4.211 | 4.16 | 4.491 | 0.876 | 0.764 | 0.981 |
| LUSC | STK11 | 2185 | 41 | 1.876 | 1.567 | 3.593 | 0.405 | 0.015 | 0.076 |
| LUSC | KEAP1 | 2185 | 248 | 11.35 | 12.048 | 7.485 | 1.362 | 0.197 | 0.342 |
| LUSC | SMARCA4 | 2185 | 108 | 4.943 | 5.078 | 4.192 | 0.99 | 0.908 | 0.981 |
| LUSC | RB1 | 2183 | 157 | 7.192 | 8.167 | 1.796 | 4.03 | <0.001 | 0.006 |
| LUSC | NF1 | 2183 | 202 | 9.253 | 9.681 | 6.886 | 1.163 | 0.589 | 0.807 |
| LUSC | BRAF | 2183 | 52 | 2.382 | 2.434 | 2.096 | 0.859 | 0.886 | 0.981 |
| LUSC | PIK3CA | 2181 | 272 | 12.471 | 12.392 | 12.913 | 0.736 | 0.116 | 0.273 |
| LUSC | MET | 2179 | 41 | 1.882 | 1.95 | 1.502 | 1.168 | 0.943 | 0.981 |
| LUSC | ERBB2 | 2174 | 38 | 1.748 | 1.9 | 0.904 | 1.62 | 0.564 | 0.807 |
| LUSC | ATM | 2183 | 107 | 4.902 | 4.759 | 5.689 | 0.625 | 0.107 | 0.273 |
| LUSC | ARID1A | 2183 | 195 | 8.933 | 9.194 | 7.485 | 0.937 | 0.865 | 0.981 |
| LUSC | NFE2L2 | 2174 | 345 | 15.869 | 16.965 | 9.726 | 1.491 | 0.054 | 0.18 |
| LUSC | PTEN | 2178 | 210 | 9.642 | 10.515 | 4.805 | 1.928 | 0.017 | 0.076 |
| LUSC | KMT2D | 2178 | 495 | 22.727 | 24.173 | 14.715 | 1.367 | 0.075 | 0.216 |
| LUSC | CDKN2A | 2185 | 425 | 19.451 | 21.664 | 7.186 | 2.766 | <0.001 | <0.001 |
| LUSC | APC | 2185 | 78 | 3.57 | 3.836 | 2.096 | 1.469 | 0.439 | 0.672 |
| LUSC | NOTCH1 | 2168 | 207 | 9.548 | 10.217 | 5.793 | 1.477 | 0.151 | 0.28 |
| LUSC | CREBBP | 2176 | 158 | 7.261 | 6.833 | 9.639 | 0.512 | 0.002 | 0.013 |
| LUSC | EP300 | 2174 | 100 | 4.6 | 4.723 | 3.916 | 0.956 | 0.992 | 0.992 |
| LUSC | PTPRD | 1796 | 132 | 7.35 | 8.055 | 3.346 | 2.024 | 0.055 | 0.18 |
| LUSC | SETD2 | 2180 | 84 | 3.853 | 4.277 | 1.502 | 2.112 | 0.148 | 0.28 |
| LUSC | CTNNB1 | 2175 | 24 | 1.103 | 0.975 | 1.824 | 0.431 | 0.136 | 0.28 |
| LUSC | MGA | 1790 | 88 | 4.916 | 5.049 | 4.151 | 1.027 | 0.931 | 0.981 |
| LUSC | CIC | 2174 | 42 | 1.932 | 2.117 | 0.904 | 1.927 | 0.403 | 0.654 |

**Supplementary Data File D3 (continued).** Co-alteration with TP53 by driver group.

| group | gene | n | n alt | % | CMH OR | p | q |
| --- | --- | --- | --- | --- | --- | --- | --- |
| EGFR-activating | KRAS | 3239 | 15 | 0.463 | 0.359 | 0.093 | 0.268 |
| EGFR-activating | STK11 | 3239 | 26 | 0.803 | 1.811 | 0.246 | 0.537 |
| EGFR-activating | KEAP1 | 3239 | 22 | 0.679 | 1.171 | 0.887 | 0.989 |
| EGFR-activating | SMARCA4 | 3239 | 87 | 2.686 | 1.04 | 0.951 | 0.989 |
| EGFR-activating | RB1 | 3239 | 211 | 6.514 | 4.958 | <0.001 | <0.001 |
| EGFR-activating | NF1 | 3239 | 61 | 1.883 | 1.039 | 0.989 | 0.989 |
| EGFR-activating | BRAF | 3239 | 32 | 0.988 | 0.626 | 0.245 | 0.537 |
| EGFR-activating | PIK3CA | 3239 | 223 | 6.885 | 0.668 | 0.006 | 0.023 |
| EGFR-activating | MET | 3239 | 23 | 0.71 | 0.765 | 0.665 | 0.989 |
| EGFR-activating | ERBB2 | 3239 | 35 | 1.081 | 1.097 | 0.922 | 0.989 |
| EGFR-activating | ATM | 3239 | 82 | 2.532 | 0.482 | 0.002 | 0.009 |
| EGFR-activating | ARID1A | 3239 | 90 | 2.779 | 1.029 | 0.986 | 0.989 |
| EGFR-activating | NFE2L2 | 3239 | 13 | 0.401 | 0.896 | 0.916 | 0.989 |
| EGFR-activating | PTEN | 3239 | 122 | 3.767 | 0.975 | 0.966 | 0.989 |
| EGFR-activating | KMT2D | 3239 | 73 | 2.254 | 0.803 | 0.428 | 0.82 |
| EGFR-activating | CDKN2A | 3239 | 82 | 2.532 | 1.332 | 0.257 | 0.537 |
| EGFR-activating | SETD2 | 3239 | 77 | 2.377 | 0.391 | <0.001 | <0.001 |
| EGFR-activating | CTNNB1 | 3239 | 192 | 5.928 | 0.387 | <0.001 | <0.001 |
| EGFR-activating | U2AF1 | 3239 | 41 | 1.266 | 0.191 | <0.001 | <0.001 |
| EGFR-activating | EP300 | 3239 | 31 | 0.957 | 1.034 | 0.922 | 0.989 |
| EGFR-activating | NOTCH1 | 3239 | 40 | 1.235 | 1.169 | 0.761 | 0.989 |
| EGFR-activating | PTPRD | 3239 | 62 | 1.914 | 1.753 | 0.064 | 0.212 |
| EGFR-activating | CIC | 3239 | 22 | 0.679 | 1.484 | 0.526 | 0.931 |
| EGFR/ALK wild-type | KRAS | 7160 | 2974 | 41.536 | 0.401 | <0.001 | <0.001 |
| EGFR/ALK wild-type | STK11 | 7160 | 1402 | 19.581 | 0.404 | <0.001 | <0.001 |
| EGFR/ALK wild-type | KEAP1 | 7160 | 1206 | 16.844 | 0.688 | <0.001 | <0.001 |
| EGFR/ALK wild-type | SMARCA4 | 7160 | 674 | 9.413 | 0.998 | 0.988 | 0.988 |
| EGFR/ALK wild-type | RB1 | 7160 | 302 | 4.218 | 5.16 | <0.001 | <0.001 |
| EGFR/ALK wild-type | NF1 | 7160 | 707 | 9.874 | 1.994 | <0.001 | <0.001 |
| EGFR/ALK wild-type | BRAF | 7160 | 519 | 7.249 | 1.031 | 0.796 | 0.832 |
| EGFR/ALK wild-type | PIK3CA | 7160 | 344 | 4.805 | 1.2 | 0.141 | 0.18 |
| EGFR/ALK wild-type | MET | 7160 | 374 | 5.224 | 1.121 | 0.356 | 0.431 |
| EGFR/ALK wild-type | ERBB2 | 7160 | 379 | 5.293 | 1.558 | <0.001 | <0.001 |
| EGFR/ALK wild-type | ATM | 7160 | 701 | 9.791 | 0.36 | <0.001 | <0.001 |
| EGFR/ALK wild-type | ARID1A | 7160 | 512 | 7.151 | 1.574 | <0.001 | <0.001 |
| EGFR/ALK wild-type | NFE2L2 | 7160 | 132 | 1.844 | 1.148 | 0.502 | 0.55 |
| EGFR/ALK wild-type | PTEN | 7160 | 127 | 1.774 | 1.703 | 0.009 | 0.014 |
| EGFR/ALK wild-type | KMT2D | 7160 | 509 | 7.109 | 1.467 | <0.001 | <0.001 |
| EGFR/ALK wild-type | CDKN2A | 7160 | 408 | 5.698 | 2.765 | <0.001 | <0.001 |
| EGFR/ALK wild-type | SETD2 | 7160 | 466 | 6.508 | 0.821 | 0.062 | 0.084 |
| EGFR/ALK wild-type | CTNNB1 | 7160 | 190 | 2.654 | 0.684 | 0.019 | 0.027 |
| EGFR/ALK wild-type | U2AF1 | 7160 | 179 | 2.5 | 0.872 | 0.443 | 0.51 |
| EGFR/ALK wild-type | EP300 | 7160 | 205 | 2.863 | 1.591 | 0.004 | 0.007 |
| EGFR/ALK wild-type | NOTCH1 | 7160 | 270 | 3.771 | 1.878 | <0.001 | <0.001 |
| EGFR/ALK wild-type | PTPRD | 7160 | 758 | 10.587 | 1.518 | <0.001 | <0.001 |
| EGFR/ALK wild-type | CIC | 7160 | 169 | 2.36 | 1.594 | 0.007 | 0.011 |

**Supplementary Data File D4.**
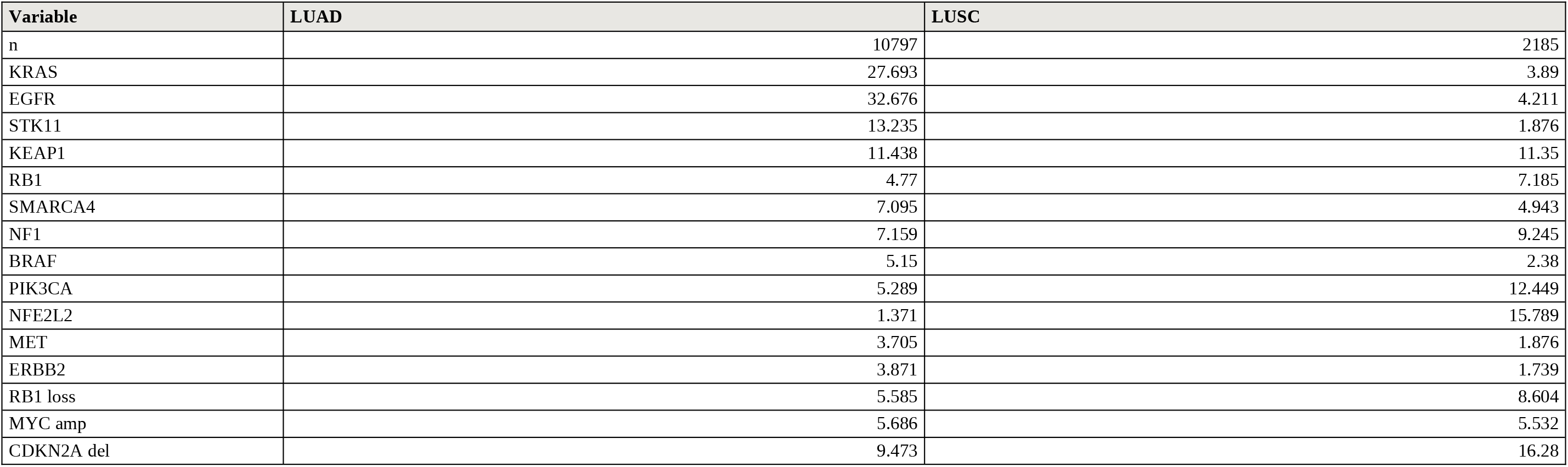
Driver and co-alteration prevalence by histology.

**Supplementary Data File D4 (continued).**
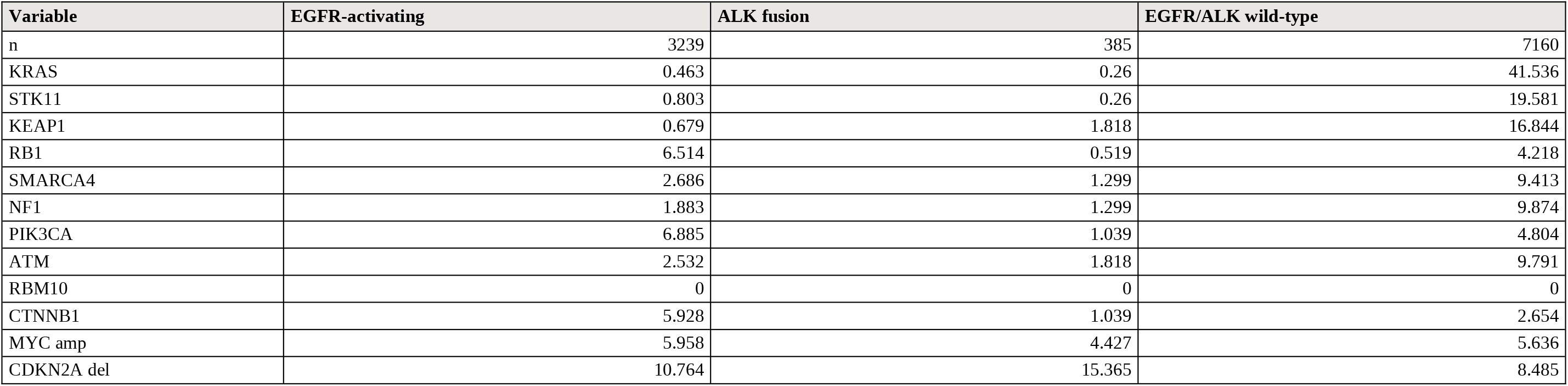
Co-alteration prevalence by driver group.

| Variable | EGFR-activating | ALK fusion | EGFR/ALK wild-type |
| --- | --- | --- | --- |
| n | 3239 | 385 | 7160 |
| KRAS | 0.463 | 0.26 | 41.536 |
| STK11 | 0.803 | 0.26 | 19.581 |
| KEAP1 | 0.679 | 1.818 | 16.844 |
| RB1 | 6.514 | 0.519 | 4.218 |
| SMARCA4 | 2.686 | 1.299 | 9.413 |
| NF1 | 1.883 | 1.299 | 9.874 |
| PIK3CA | 6.885 | 1.039 | 4.804 |
| ATM | 2.532 | 1.818 | 9.791 |
| RBM10 | 0 | 0 | 0 |
| CTNNB1 | 5.928 | 1.039 | 2.654 |
| MYC amp | 5.958 | 4.427 | 5.636 |
| CDKN2A del | 10.764 | 15.365 | 8.485 |

**Supplementary Data File D5.**
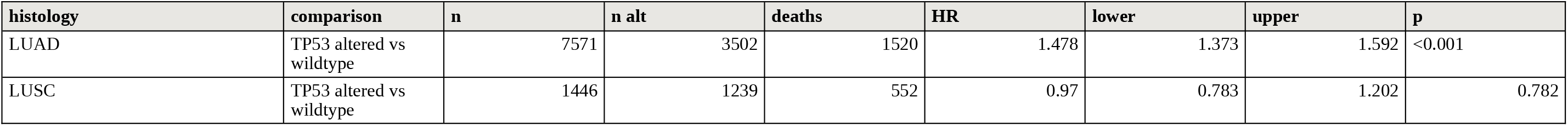
TP53 survival estimates by histology, validation cohort.

**Supplementary Data File D5 (continued).** TP53 survival estimates by driver group.

| group | comparison | n | n alt | deaths | HR | lower | upper | p |
| --- | --- | --- | --- | --- | --- | --- | --- | --- |
| all LUAD | EGFR-activating vs EGFR/ALK-WT | 7571 | 2063 | 653 | 0.687 | 0.628 | 0.751 | <0.001 |
| all LUAD | ALK fusion vs EGFR/ALK-WT | 7571 | 244 | 65 | 0.545 | 0.425 | 0.698 | <0.001 |
| EGFR-activating | TP53 altered vs wildtype | 2061 | 1097 | 453 | 2.272 | 1.919 | 2.689 | <0.001 |
| ALK fusion | TP53 altered vs wildtype | 240 | 54 | 24 | 2.415 | 1.421 | 4.105 | 0.001 |
| EGFR/ALK wild-type | TP53 altered vs wildtype | 5268 | 2349 | 1042 | 1.305 | 1.197 | 1.422 | <0.001 |

**Supplementary Data File D6.** Mutually adjusted survival models by histology.

| histology | term | n | n alt | deaths | HR | lower | upper | p | q |
| --- | --- | --- | --- | --- | --- | --- | --- | --- | --- |
| LUAD | TP53_alt | 7314 | 3374 | 1457 | 1.531 | 1.411 | 1.661 | <0.001 | <0.001 |
| LUAD | KRAS_mut | 7314 | 2281 | 873 | 1.096 | 1.001 | 1.201 | 0.047 | 0.078 |
| LUAD | EGFR_mut | 7314 | 2172 | 707 | 0.792 | 0.716 | 0.875 | <0.001 | <0.001 |
| LUAD | STK11_mut | 7314 | 1049 | 510 | 1.309 | 1.171 | 1.463 | <0.001 | <0.001 |
| LUAD | KEAP1_mut | 7314 | 947 | 489 | 1.458 | 1.304 | 1.63 | <0.001 | <0.001 |
| LUAD | SMARCA4_mut | 7314 | 518 | 277 | 1.417 | 1.247 | 1.611 | <0.001 | <0.001 |
| LUAD | RB1_loss | 7314 | 399 | 180 | 1.152 | 0.985 | 1.348 | 0.077 | 0.099 |
| LUAD | NF1_mut | 7314 | 553 | 204 | 0.839 | 0.724 | 0.972 | 0.019 | 0.039 |
| LUAD | BRAF_mut | 7314 | 408 | 161 | 0.989 | 0.841 | 1.164 | 0.898 | 0.91 |
| LUAD | PIK3CA_mut | 7314 | 386 | 158 | 0.984 | 0.837 | 1.158 | 0.85 | 0.91 |
| LUAD | NFE2L2_mut | 7314 | 110 | 59 | 1.419 | 1.091 | 1.846 | 0.009 | 0.021 |
| LUAD | ATM_mut | 7314 | 532 | 210 | 1.008 | 0.873 | 1.165 | 0.91 | 0.91 |
| LUAD | ARID1A_mut | 7314 | 424 | 191 | 1.165 | 1.002 | 1.354 | 0.048 | 0.078 |
| LUAD | KMT2D_mut | 7314 | 421 | 173 | 0.917 | 0.783 | 1.075 | 0.285 | 0.341 |
| LUAD | MYC_amp | 7314 | 423 | 205 | 1.466 | 1.267 | 1.696 | <0.001 | <0.001 |
| LUAD | CCNE1_amp | 7314 | 182 | 79 | 1.242 | 0.988 | 1.562 | 0.063 | 0.091 |
| LUAD | MDM2_amp | 7314 | 441 | 156 | 1.17 | 0.99 | 1.384 | 0.066 | 0.091 |
| LUAD | CDKN2A_del | 7314 | 756 | 381 | 1.738 | 1.554 | 1.945 | <0.001 | <0.001 |
| LUSC | TP53_alt | 1407 | 1205 | 522 | 0.932 | 0.743 | 1.17 | 0.544 | 0.818 |
| LUSC | KRAS_mut | 1407 | 41 | 24 | 1.601 | 1.053 | 2.435 | 0.028 | 0.368 |
| LUSC | EGFR_mut | 1407 | 60 | 27 | 1.017 | 0.686 | 1.507 | 0.933 | 0.933 |
| LUSC | STK11_mut | 1407 | 28 | 16 | 1.709 | 1.022 | 2.857 | 0.041 | 0.368 |
| LUSC | KEAP1_mut | 1407 | 166 | 83 | 1.255 | 0.985 | 1.598 | 0.066 | 0.396 |
| LUSC | SMARCA4_mut | 1407 | 67 | 27 | 0.88 | 0.591 | 1.309 | 0.527 | 0.818 |
| LUSC | RB1_loss | 1407 | 137 | 54 | 0.841 | 0.633 | 1.118 | 0.233 | 0.705 |
| LUSC | NF1_mut | 1407 | 141 | 67 | 1.033 | 0.793 | 1.345 | 0.811 | 0.858 |
| LUSC | BRAF_mut | 1407 | 36 | 13 | 0.714 | 0.41 | 1.245 | 0.235 | 0.705 |
| LUSC | PIK3CA_mut | 1407 | 166 | 68 | 0.883 | 0.682 | 1.143 | 0.346 | 0.778 |
| LUSC | NFE2L2_mut | 1407 | 218 | 96 | 1.131 | 0.903 | 1.417 | 0.286 | 0.735 |
| LUSC | ATM_mut | 1407 | 76 | 33 | 1.067 | 0.74 | 1.539 | 0.727 | 0.818 |
| LUSC | ARID1A_mut | 1407 | 128 | 61 | 1.05 | 0.799 | 1.38 | 0.727 | 0.818 |
| LUSC | KMT2D_mut | 1407 | 335 | 146 | 1.134 | 0.936 | 1.375 | 0.2 | 0.705 |
| LUSC | MYC_amp | 1407 | 79 | 36 | 1.067 | 0.747 | 1.523 | 0.722 | 0.818 |
| LUSC | CCNE1_amp | 1407 | 60 | 21 | 0.873 | 0.556 | 1.37 | 0.554 | 0.818 |
| LUSC | MDM2_amp | 1407 | 39 | 15 | 0.823 | 0.49 | 1.385 | 0.464 | 0.818 |
| LUSC | CDKN2A_del | 1407 | 255 | 107 | 1.043 | 0.841 | 1.293 | 0.705 | 0.818 |

**Supplementary Data File D6 (continued).** Mutually adjusted survival models by driver group.

| group | term | n | n alt | deaths | HR | lower | upper | p | q |
| --- | --- | --- | --- | --- | --- | --- | --- | --- | --- |
| EGFR-activating | TP53_alt | 1979 | 1055 | 433 | 2.242 | 1.859 | 2.705 | <0.001 | <0.001 |
| EGFR-activating | KRAS_mut | 1979 | 13 | 7 | 1.229 | 0.547 | 2.763 | 0.618 | 0.618 |
| EGFR-activating | STK11_mut | 1979 | 18 | 9 | 1.942 | 0.989 | 3.814 | 0.054 | 0.086 |
| EGFR-activating | SMARCA4_mut | 1979 | 55 | 27 | 1.498 | 1.003 | 2.237 | 0.048 | 0.086 |
| EGFR-activating | RB1_loss | 1979 | 164 | 76 | 1.311 | 1.018 | 1.689 | 0.036 | 0.086 |
| EGFR-activating | NF1_mut | 1979 | 35 | 14 | 1.185 | 0.682 | 2.059 | 0.548 | 0.584 |
| EGFR-activating | BRAF_mut | 1979 | 25 | 10 | 1.354 | 0.705 | 2.603 | 0.363 | 0.446 |
| EGFR-activating | PIK3CA_mut | 1979 | 132 | 56 | 1.451 | 1.092 | 1.928 | 0.01 | 0.042 |
| EGFR-activating | ATM_mut | 1979 | 51 | 20 | 1.608 | 1.013 | 2.552 | 0.044 | 0.086 |
| EGFR-activating | ARID1A_mut | 1979 | 56 | 31 | 1.898 | 1.279 | 2.815 | 0.002 | 0.008 |
| EGFR-activating | KMT2D_mut | 1979 | 42 | 14 | 1.186 | 0.692 | 2.031 | 0.536 | 0.584 |
| EGFR-activating | CTNNB1_mut | 1979 | 119 | 28 | 0.695 | 0.471 | 1.027 | 0.068 | 0.098 |
| EGFR-activating | MYC_amp | 1979 | 128 | 51 | 1.427 | 1.063 | 1.917 | 0.018 | 0.058 |
| EGFR-activating | CCNE1_amp | 1979 | 78 | 34 | 1.456 | 1.021 | 2.076 | 0.038 | 0.086 |
| EGFR-activating | MDM2_amp | 1979 | 175 | 50 | 1.23 | 0.901 | 1.68 | 0.193 | 0.257 |
| EGFR-activating | CDKN2A_del | 1979 | 263 | 110 | 1.618 | 1.304 | 2.008 | <0.001 | <0.001 |
| EGFR/ALK wild-type | TP53_alt | 5094 | 2264 | 1000 | 1.323 | 1.204 | 1.455 | <0.001 | <0.001 |
| EGFR/ALK wild-type | KRAS_mut | 5094 | 2268 | 866 | 1.023 | 0.932 | 1.124 | 0.628 | 0.707 |
| EGFR/ALK wild-type | STK11_mut | 5094 | 1030 | 501 | 1.231 | 1.098 | 1.38 | <0.001 | 0.001 |
| EGFR/ALK wild-type | KEAP1_mut | 5094 | 931 | 480 | 1.451 | 1.294 | 1.626 | <0.001 | <0.001 |
| EGFR/ALK wild-type | SMARCA4_mut | 5094 | 459 | 249 | 1.417 | 1.236 | 1.624 | <0.001 | <0.001 |
| EGFR/ALK wild-type | RB1_loss | 5094 | 232 | 103 | 1.03 | 0.839 | 1.265 | 0.775 | 0.81 |
| EGFR/ALK wild-type | NF1_mut | 5094 | 514 | 189 | 0.833 | 0.714 | 0.972 | 0.021 | 0.046 |
| EGFR/ALK wild-type | BRAF_mut | 5094 | 379 | 150 | 0.942 | 0.795 | 1.117 | 0.493 | 0.592 |
| EGFR/ALK wild-type | PIK3CA_mut | 5094 | 251 | 102 | 0.879 | 0.718 | 1.075 | 0.208 | 0.374 |
| EGFR/ALK wild-type | NFE2L2_mut | 5094 | 100 | 57 | 1.562 | 1.194 | 2.044 | 0.001 | 0.003 |
| EGFR/ALK wild-type | ATM_mut | 5094 | 474 | 187 | 0.928 | 0.796 | 1.082 | 0.339 | 0.474 |
| EGFR/ALK wild-type | ARID1A_mut | 5094 | 361 | 157 | 1.079 | 0.914 | 1.275 | 0.369 | 0.474 |
| EGFR/ALK wild-type | KMT2D_mut | 5094 | 370 | 154 | 0.904 | 0.763 | 1.071 | 0.243 | 0.397 |
| EGFR/ALK wild-type | CTNNB1_mut | 5094 | 133 | 53 | 1.034 | 0.786 | 1.362 | 0.81 | 0.81 |
| EGFR/ALK wild-type | MYC_amp | 5094 | 284 | 153 | 1.53 | 1.291 | 1.814 | <0.001 | <0.001 |
| EGFR/ALK wild-type | CCNE1_amp | 5094 | 103 | 45 | 1.149 | 0.849 | 1.555 | 0.368 | 0.474 |
| EGFR/ALK wild-type | MDM2_amp | 5094 | 250 | 102 | 1.205 | 0.981 | 1.481 | 0.076 | 0.152 |
| EGFR/ALK wild-type | CDKN2A_del | 5094 | 447 | 255 | 1.796 | 1.566 | 2.059 | <0.001 | <0.001 |

**Supplementary Data File D7.** Discovery cohort characteristics by histology.

| characteristic | level | Adenocarcinoma (n=2,797) | Squamous (n=498) | p |
| --- | --- | --- | --- | --- |
| Age (years) | median | 69 | 73 | <0.001 |
| Age (years) | range | 23–96 | 43–93 |  |
| Age (years) | SD | 11.18 | 9.38 |  |
| Race | White | 1351 (51.0%) | 313 (64.9%) | <0.001 |
| Race | Asian | 736 (27.8%) | 70 (14.5%) | <0.001 |
| Race | Black | 222 (8.4%) | 55 (11.4%) | <0.001 |
| Race | Hispanic | 113 (4.3%) | 17 (3.5%) | <0.001 |
| Race | Other | 229 (8.6%) | 27 (5.6%) | <0.001 |
| Sex | Female | 1640 (58.6%) | 213 (42.8%) | <0.001 |
| Sex | Male | 1157 (41.4%) | 285 (57.2%) | <0.001 |
| Charlson index | median | 1 | 2 | <0.001 |
| Charlson index | range | 0–14 | 0–11 |  |
| Performance status | ECOG 0–1 | 218 (7.8%) | 37 (7.4%) | 0.242 |
| Performance status | ECOG 2–4 | 31 (1.1%) | 10 (2.0%) | 0.242 |
| Performance status | unknown | 2548 (91.1%) | 451 (90.6%) | 0.242 |
| TP53 | wild-type | 1361 (48.7%) | 106 (21.3%) | <0.001 |
| EGFR | pathogenic variant | 852 (30.5%) | 35 (7.0%) | <0.001 |
| KRAS | pathogenic variant | 805 (28.8%) | 36 (7.2%) | <0.001 |
| CDKN2A | pathogenic variant | 576 (20.6%) | 179 (35.9%) | <0.001 |
| PIK3CA | pathogenic variant | 136 (4.9%) | 117 (23.5%) | <0.001 |
| MET | pathogenic variant | 167 (6.0%) | 15 (3.0%) | 0.008 |
| RB1 | pathogenic variant | 155 (5.5%) | 38 (7.6%) | 0.067 |
| MDM2 | amplification | 140 (5.0%) | 17 (3.4%) | 0.124 |
| BRAF | pathogenic variant | 138 (4.9%) | 7 (1.4%) | <0.001 |
| MYC | amplification | 123 (4.4%) | 18 (3.6%) | 0.426 |
| STK11 | pathogenic variant | 128 (4.6%) | 5 (1.0%) | <0.001 |
| ALK | pathogenic variant | 131 (4.7%) | 2 (0.4%) | <0.001 |
| ROS1 | pathogenic variant | 34 (1.2%) | 0 (0.0%) | 0.013 |
| ERBB2 | pathogenic variant | 102 (3.6%) | 7 (1.4%) | 0.01 |
| KEAP1 | pathogenic variant | 86 (3.1%) | 17 (3.4%) | 0.689 |
| NF1 | pathogenic variant | 73 (2.6%) | 18 (3.6%) | 0.207 |
| ATM | pathogenic variant | 85 (3.0%) | 7 (1.4%) | 0.041 |
| SMARCA4 | pathogenic variant | 53 (1.9%) | 7 (1.4%) | 0.452 |
| BRCA2 | pathogenic variant | 41 (1.5%) | 12 (2.4%) | 0.123 |

**Supplementary Data File D7 (continued).** Discovery cohort characteristics by driver status.

| characteristic | level | EGFR/ALK/ROS1 positive (n=1,053) | negative (n=2,242) | p |
| --- | --- | --- | --- | --- |
| Age (years) | median | 67 | 71 | <0.001 |
| Age (years) | SD | 12.64 | 9.77 | <0.001 |
| Race | White | 358 (35.6%) | 1306 (61.4%) | <0.001 |
| Race | Asian | 457 (45.5%) | 349 (16.4%) | <0.001 |
| Race | Black | 42 (4.2%) | 235 (11.0%) | <0.001 |
| Race | Hispanic | 54 (5.4%) | 76 (3.6%) | <0.001 |
| Race | Other | 94 (9.4%) | 162 (7.6%) | <0.001 |
| Sex | Female | 690 (65.5%) | 1163 (51.9%) | <0.001 |
| Sex | Male | 363 (34.5%) | 1079 (48.1%) | <0.001 |
| Charlson index | median | 1 | 2 | <0.001 |
| Performance status | ECOG 0–1 | 74 (7.0%) | 181 (8.1%) | 0.322 |
| Performance status | ECOG 2–4 | 10 (0.9%) | 31 (1.4%) | 0.322 |
| TP53 | wild-type | 492 (46.7%) | 975 (43.5%) | 0.136 |
| KRAS | pathogenic variant | 21 (2.0%) | 820 (36.6%) | <0.001 |
| STK11 | pathogenic variant | 0 (0.0%) | 133 (5.9%) | <0.001 |
| CDKN2A | pathogenic variant | 268 (25.5%) | 487 (21.7%) | 0.017 |
| MDM2 | amplification | 82 (7.8%) | 75 (3.3%) | <0.001 |
| MYC | amplification | 47 (4.5%) | 94 (4.2%) | 0.72 |
| RB1 | pathogenic variant | 67 (6.4%) | 126 (5.6%) | 0.397 |
| MET | pathogenic variant | 33 (3.1%) | 149 (6.6%) | <0.001 |
| BRAF | pathogenic variant | 12 (1.1%) | 133 (5.9%) | <0.001 |
| PIK3CA | pathogenic variant | 64 (6.1%) | 189 (8.4%) | 0.018 |
| CTNNB1 | pathogenic variant | 77 (7.3%) | 45 (2.0%) | <0.001 |
| ERBB2 | pathogenic variant | 12 (1.1%) | 97 (4.3%) | <0.001 |

**Supplementary Data File D8.** Validation cohort demographics by histology.

| Variable | LUAD | LUSC | All lung |
| --- | --- | --- | --- |
| n | 10797 | 2185 | 13827 |
| age known % | 90.4 | 98.3 | 92 |
| age median | 66.2 | 68 | 66.5 |
| age q1 | 58.4 | 61.1 | 59 |
| age q3 | 73.8 | 74.5 | 73.8 |
| sex known % | 96.7 | 99.9 | 97.3 |
| female % | 58.5 | 28.6 | 52.6 |
| race known % | 61.7 | 44.6 | 57.5 |
| White % | 82.2 | 85.7 | 82.9 |
| Asian % | 12.4 | 8.3 | 11.7 |
| Black % | 5.4 | 6 | 5.4 |
| ethnicity known % | 24.2 | 18.6 | 22 |
| Hispanic % | 11.1 | 17 | 11.8 |
| smoking known % | 32.3 | 45.4 | 32.5 |
| never smoker % | 30.1 | 3.9 | 24.4 |
| stage known % | 49.3 | 60 | 49.3 |
| stage IV % | 30 | 14.9 | 27.3 |
| sampletype known % | 80.8 | 71.7 | 80.3 |
| metastasis % | 35.4 | 20.7 | 33.4 |
| whole exome % | 17.6 | 38.4 | 21.1 |
| studies | 34 | 17 | 41 |

**Supplementary Data File D8 (continued).** Validation adenocarcinoma demographics by driver group.

| Variable | EGFR-activating | ALK fusion | EGFR/ALK wild-type | All adenocarcinoma |
| --- | --- | --- | --- | --- |
| n | 3239 | 385 | 7160 | 10784 |
| age known % | 92.9 | 94.3 | 89 | 90.4 |
| age median | 64 | 58 | 68 | 66.2 |
| age q1 | 56 | 48 | 60 | 58.4 |
| age q3 | 72 | 66 | 74.8 | 73.8 |
| sex known % | 98.6 | 100 | 95.8 | 96.8 |
| female % | 67.1 | 61.3 | 54.4 | 58.5 |
| race known % | 55.4 | 59 | 64.7 | 61.7 |
| White % | 67.5 | 79.8 | 87.8 | 82.1 |
| Asian % | 26.6 | 13.3 | 7 | 12.4 |
| Black % | 5.9 | 6.9 | 5.2 | 5.4 |
| ethnicity known % | 23.5 | 20.8 | 24.7 | 24.2 |
| Hispanic % | 10.4 | 17.5 | 11.1 | 11.1 |
| smoking known % | 28.4 | 20.3 | 34.8 | 32.3 |
| never smoker % | 58.1 | 69.2 | 18.5 | 30.1 |
| stage known % | 57.2 | 51.7 | 45.6 | 49.3 |
| stage IV % | 35.3 | 42.7 | 26.2 | 30 |
| sampletype known % | 85.1 | 96.6 | 78 | 80.8 |
| metastasis % | 33.4 | 46.5 | 35.5 | 35.3 |
| whole exome % | 13.1 | 4.2 | 20.2 | 17.5 |
| studies | 25 | 11 | 31 | 31 |

**Supplementary Data File D8 (continued).** Tests of demographic difference between driver groups.

| group | variable | group % | wildtype % | group n | wildtype n | OR | p |
| --- | --- | --- | --- | --- | --- | --- | --- |
| EGFR-activating | never smoker | 58.1 | 18.5 | 919 | 2490 | 6.121 | <0.001 |
| EGFR-activating | Asian | 26.6 | 7 | 1682 | 4428 | 4.839 | <0.001 |
| EGFR-activating | female | 67.1 | 54.4 | 3193 | 6857 | 1.713 | <0.001 |
| EGFR-activating | stage IV | 35.3 | 26.2 | 1853 | 3264 | 1.538 | <0.001 |
| EGFR-activating | metastasis | 33.4 | 35.5 | 2758 | 5586 | 0.911 | 0.06 |
| EGFR-activating | age (median) | 64 | 68 | 3009 | 6374 |  | <0.001 |
| ALK fusion | never smoker | 69.2 | 18.5 | 78 | 2490 | 9.929 | <0.001 |
| ALK fusion | Asian | 13.3 | 7 | 203 | 4428 | 2.045 | 0.002 |
| ALK fusion | female | 61.3 | 54.4 | 385 | 6857 | 1.329 | 0.008 |
| ALK fusion | stage IV | 42.7 | 26.2 | 199 | 3264 | 2.097 | <0.001 |
| ALK fusion | metastasis | 46.5 | 35.5 | 372 | 5586 | 1.577 | <0.001 |
| ALK fusion | age (median) | 58 | 68 | 363 | 6374 |  | <0.001 |

**Supplementary Data File D9.**
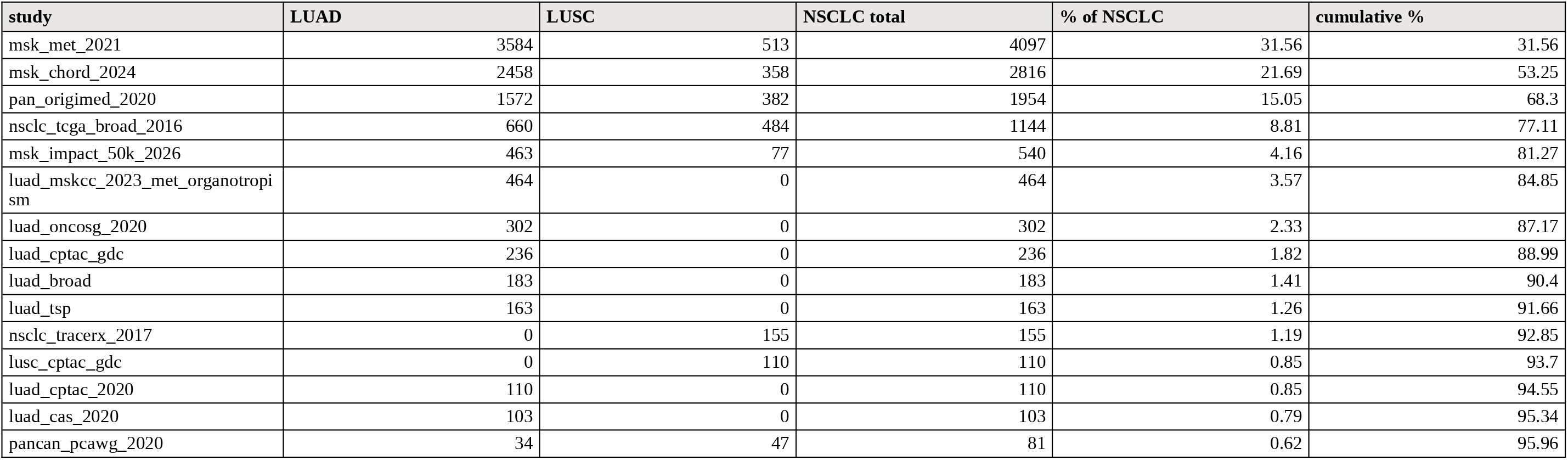
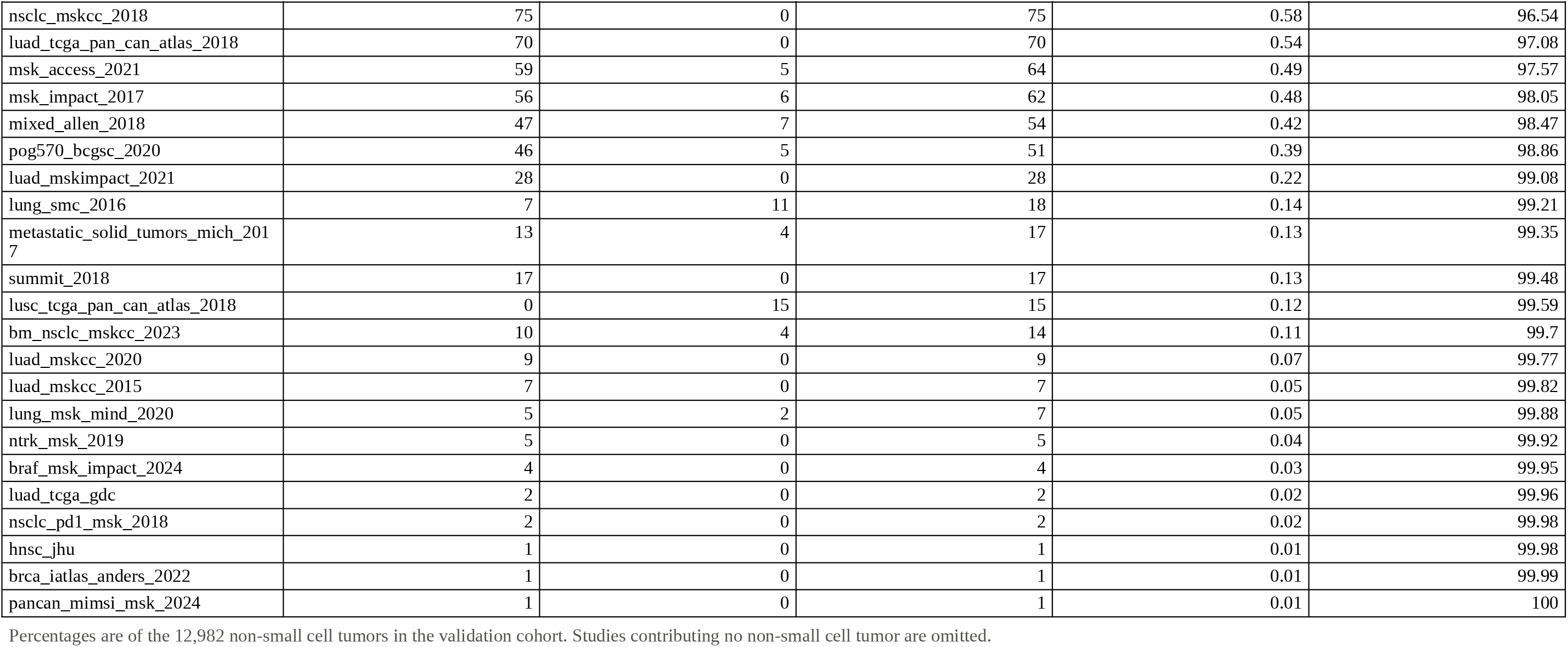
Validation cohort composition by contributing study.

**Supplementary Data File D10.**
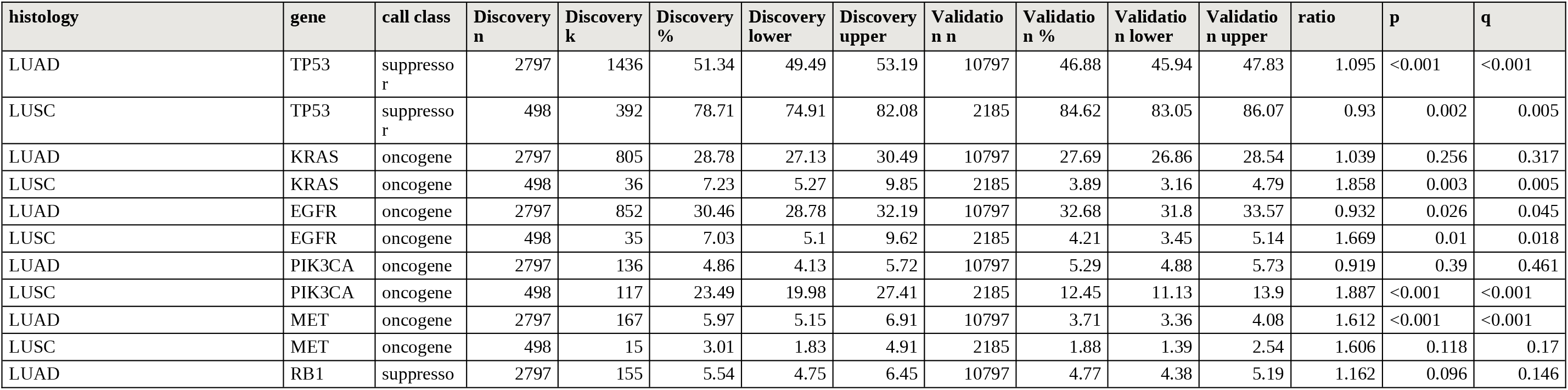
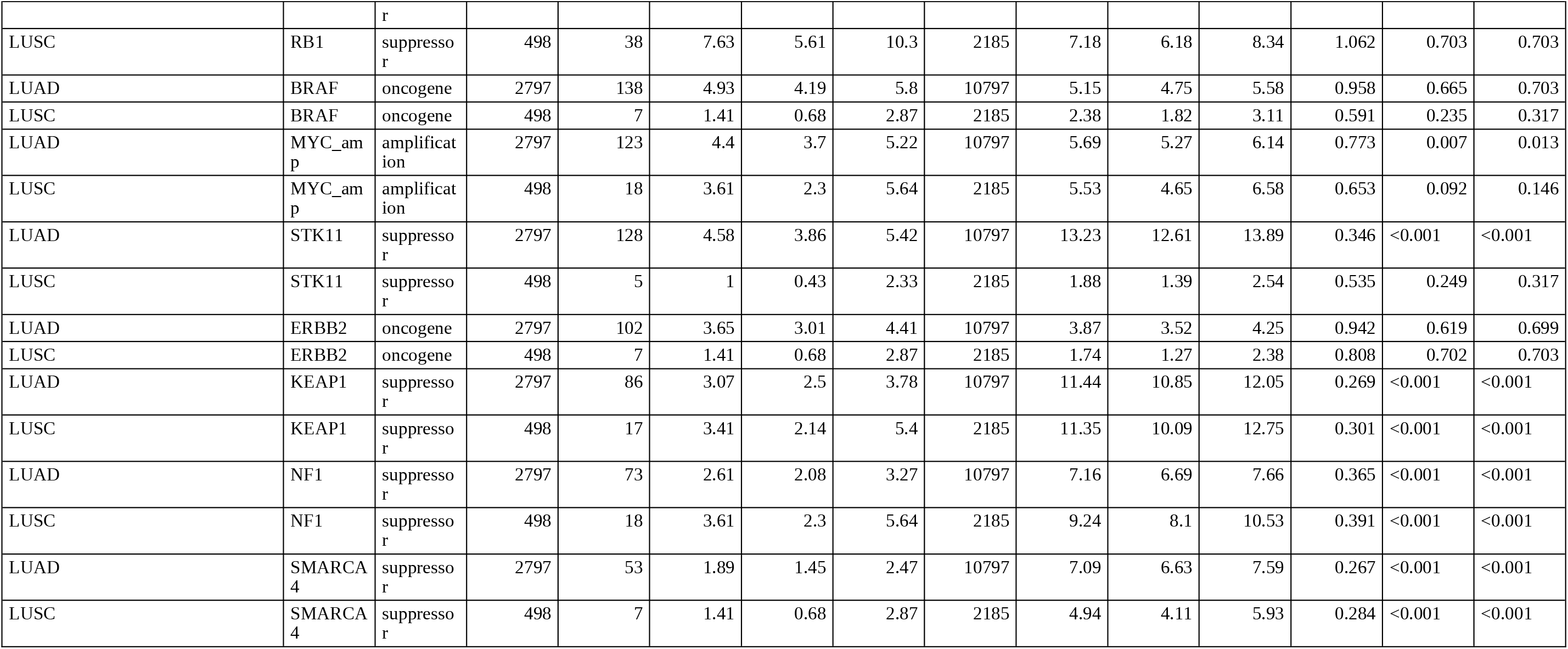
Prevalence concordance between cohorts, 26 comparisons.

**Supplementary Data File D10 (continued).** Summary by variant-call class.

| call class | n comparisons | median ratio | n concordant |
| --- | --- | --- | --- |
| amplification | 2 | 0.713 | 1 |
| oncogene | 12 | 0.999 | 7 |
| suppressor | 12 | 0.378 | 3 |

**Supplementary Data File D11.**
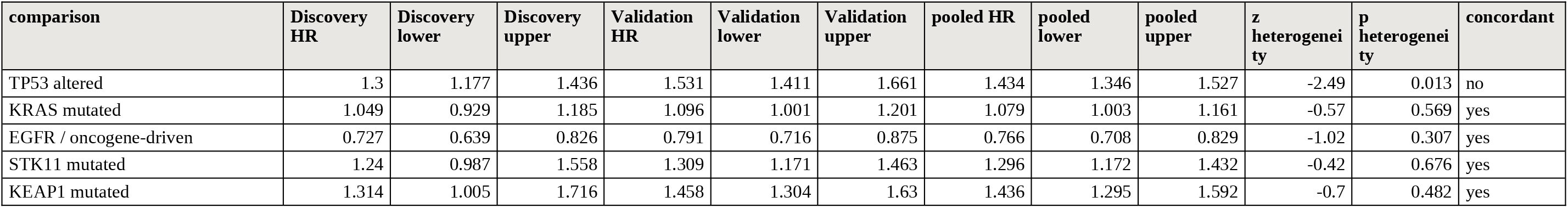
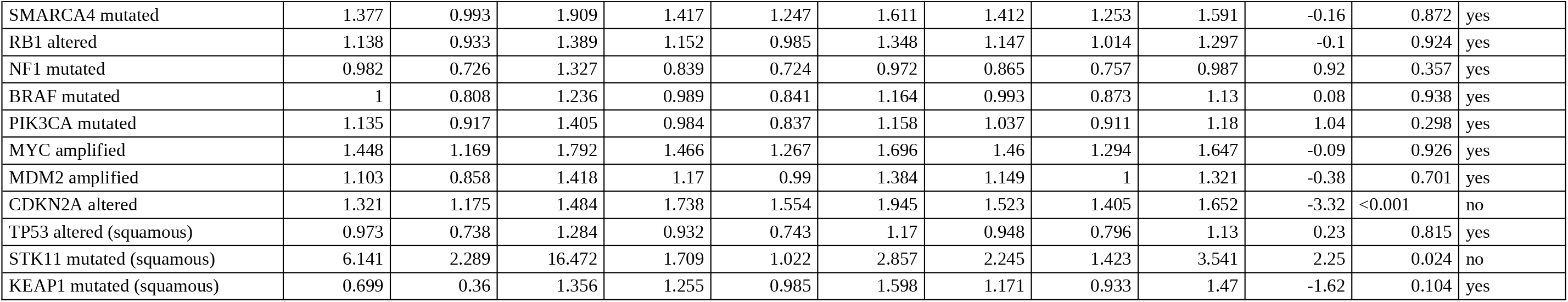
Survival-association concordance between cohorts, 16 comparisons.

| comparison | Discovery<br>HR | Discovery<br>lower | Discovery<br>upper | Validation<br>HR | Validation<br>lower | Validation<br>upper | pooled HR | pooled<br>lower | pooled<br>upper | z<br>heterogeneity | p<br>heterogeneity | concordant |
| --- | --- | --- | --- | --- | --- | --- | --- | --- | --- | --- | --- | --- |
| TP53 altered | 1.3 | 1.177 | 1.436 | 1.531 | 1.411 | 1.661 | 1.434 | 1.346 | 1.527 | -2.49 | 0.013 | no |
| KRAS mutated | 1.049 | 0.929 | 1.185 | 1.096 | 1.001 | 1.201 | 1.079 | 1.003 | 1.161 | -0.57 | 0.569 | yes |
| EGFR / oncogene-driven | 0.727 | 0.639 | 0.826 | 0.791 | 0.716 | 0.875 | 0.766 | 0.708 | 0.829 | -1.02 | 0.307 | yes |
| STK11 mutated | 1.24 | 0.987 | 1.558 | 1.309 | 1.171 | 1.463 | 1.296 | 1.172 | 1.432 | -0.42 | 0.676 | yes |
| KEAP1 mutated | 1.314 | 1.005 | 1.716 | 1.458 | 1.304 | 1.63 | 1.436 | 1.295 | 1.592 | -0.7 | 0.482 | yes |

| comparison | Discovery HR | Discovery lower | Discovery upper | Validation HR | Validation lower | Validation upper | pooled HR | pooled lower | pooled upper | z heterogeneity | p heterogeneity | concordant |
| --- | --- | --- | --- | --- | --- | --- | --- | --- | --- | --- | --- | --- |
| SMARCA4 mutated | 1.377 | 0.993 | 1.909 | 1.417 | 1.247 | 1.611 | 1.412 | 1.253 | 1.591 | -0.16 | 0.872 | yes |
| RB1 altered | 1.138 | 0.933 | 1.389 | 1.152 | 0.985 | 1.348 | 1.147 | 1.014 | 1.297 | -0.1 | 0.924 | yes |
| NF1 mutated | 0.982 | 0.726 | 1.327 | 0.839 | 0.724 | 0.972 | 0.865 | 0.757 | 0.987 | 0.92 | 0.357 | yes |
| BRAF mutated | 1 | 0.808 | 1.236 | 0.989 | 0.841 | 1.164 | 0.993 | 0.873 | 1.13 | 0.08 | 0.938 | yes |
| PIK3CA mutated | 1.135 | 0.917 | 1.405 | 0.984 | 0.837 | 1.158 | 1.037 | 0.911 | 1.18 | 1.04 | 0.298 | yes |
| MYC amplified | 1.448 | 1.169 | 1.792 | 1.466 | 1.267 | 1.696 | 1.46 | 1.294 | 1.647 | -0.09 | 0.926 | yes |
| MDM2 amplified | 1.103 | 0.858 | 1.418 | 1.17 | 0.99 | 1.384 | 1.149 | 1 | 1.321 | -0.38 | 0.701 | yes |
| CDKN2A altered | 1.321 | 1.175 | 1.484 | 1.738 | 1.554 | 1.945 | 1.523 | 1.405 | 1.652 | -3.32 | <0.001 | no |
| TP53 altered (squamous) | 0.973 | 0.738 | 1.284 | 0.932 | 0.743 | 1.17 | 0.948 | 0.796 | 1.13 | 0.23 | 0.815 | yes |
| STK11 mutated (squamous) | 6.141 | 2.289 | 16.472 | 1.709 | 1.022 | 2.857 | 2.245 | 1.423 | 3.541 | 2.25 | 0.024 | no |
| KEAP1 mutated (squamous) | 0.699 | 0.36 | 1.356 | 1.255 | 0.985 | 1.598 | 1.171 | 0.933 | 1.47 | -1.62 | 0.104 | yes |

**Supplementary Data File D12.** TP53 hazard ratio by driver status in each cohort.

| cohort | group | n | HR | lower | upper | p |
| --- | --- | --- | --- | --- | --- | --- |
| KPNC | EGFR/ALK/ROS1 positive | 1053 | 1.797 | 1.499 | 2.155 | <0.001 |
| KPNC | EGFR/ALK/ROS1 negative | 2242 | 1.084 | 0.971 | 1.21 | 0.151 |
| cBioPortal | EGFR-activating | 3239 | 2.272 | 1.919 | 2.689 | <0.001 |
| cBioPortal | ALK fusion | 385 | 2.415 | 1.421 | 4.105 | 0.001 |
| cBioPortal | EGFR/ALK wild-type | 7160 | 1.305 | 1.197 | 1.422 | <0.001 |

**Supplementary Data File D12 (continued).** Replication and pooling of the differential association.

| cohort | n | contrast | ratio | lower | upper | p | p heterogeneity |
| --- | --- | --- | --- | --- | --- | --- | --- |
| KPNC | 3295 | TP53 HR, oncogene-positive / TP53 HR, oncogene-negative | 1.658 | 1.341 | 2.05 | <0.001 |  |
| cBioPortal | 7571 | TP53 HR, EGFR-activating / TP53 HR, EGFR/ALK wild-type | 1.741 | 1.441 | 2.104 | <0.001 |  |
| fixed-effect pooled | 10866 | TP53 HR ratio, oncogene-driven vs not | 1.704 | 1.479 | 1.962 | <0.001 | 0.736 |

**Supplementary Data File D13.** Mixture estimate for the discovery squamous label.

| marker | Discovery squamous % | expected squamous % | adenocarcinoma % | implied adeno fraction | lower | upper | direction informative |
| --- | --- | --- | --- | --- | --- | --- | --- |
| TP53 | 78.71 | 84.62 | 46.88 | 15.7 | 6.7 | 25.7 | yes |
| EGFR | 7.03 | 4.21 | 32.68 | 9.9 | 3.1 | 19 | yes |
| KRAS | 7.23 | 3.89 | 27.69 | 14 | 5.8 | 25 | yes |
| PIK3CA | 23.49 | 12.45 | 5.29 | -154.2 | -208.9 | -105.2 | no |
| STK11 | 1 | 1.88 | 13.23 | -7.8 | -12.8 | 4 | no |

**Supplementary Data File D14.** TP53 by driver interaction across adjustment models.

| model | n | deaths | term | HR | lower | upper | p |
| --- | --- | --- | --- | --- | --- | --- | --- |
| 1. TP53 x driver, unadjusted | 7558 | 2827 | t53 | 1.311 | 1.203 | 1.429 | <0.001 |
| 1. TP53 x driver, unadjusted | 7558 | 2827 | drv | 0.479 | 0.416 | 0.551 | <0.001 |
| 1. TP53 x driver, unadjusted | 7558 | 2827 | ix | 1.73 | 1.448 | 2.066 | <0.001 |
| 2. + age and sex | 7333 | 2753 | t53 | 1.289 | 1.181 | 1.407 | <0.001 |
| 2. + age and sex | 7333 | 2753 | drv | 0.49 | 0.425 | 0.565 | <0.001 |
| 2. + age and sex | 7333 | 2753 | ix | 1.766 | 1.475 | 2.113 | <0.001 |
| 3. smoking subset, no smoking term | 3120 | 1005 | t53 | 1.438 | 1.244 | 1.662 | <0.001 |
| 3. smoking subset, no smoking term | 3120 | 1005 | drv | 0.53 | 0.415 | 0.677 | <0.001 |
| 3. smoking subset, no smoking term | 3120 | 1005 | ix | 1.661 | 1.228 | 2.247 | <0.001 |
| 4. smoking subset, + smoking | 3120 | 1005 | t53 | 1.448 | 1.252 | 1.675 | <0.001 |
| 4. smoking subset, + smoking | 3120 | 1005 | drv | 0.515 | 0.4 | 0.662 | <0.001 |
| 4. smoking subset, + smoking | 3120 | 1005 | ix | 1.658 | 1.226 | 2.242 | 0.001 |
| 5. + smoking, age, sex | 3067 | 992 | t53 | 1.381 | 1.192 | 1.6 | <0.001 |
| 5. + smoking, age, sex | 3067 | 992 | drv | 0.509 | 0.395 | 0.655 | <0.001 |
| 5. + smoking, age, sex | 3067 | 992 | ix | 1.715 | 1.266 | 2.323 | <0.001 |
| 6. stage subset, + stage | 3332 | 1087 | t53 | 1.238 | 1.075 | 1.424 | 0.003 |
| 6. stage subset, + stage | 3332 | 1087 | drv | 0.466 | 0.372 | 0.582 | <0.001 |
| 6. stage subset, + stage | 3332 | 1087 | ix | 1.57 | 1.184 | 2.08 | 0.002 |

**Supplementary Data File D14 (continued).** TP53 hazard ratio within each smoking and driver stratum.

| smoking | driver | n | deaths | n TP53 | HR | lower | upper | p |
| --- | --- | --- | --- | --- | --- | --- | --- | --- |
| never smokers | oncogene-driven | 550 | 150 | 267 | 2.75 | 1.934 | 3.911 | <0.001 |
| never smokers | not driven | 396 | 141 | 139 | 1.596 | 1.128 | 2.258 | 0.008 |
| ever smokers | oncogene-driven | 375 | 113 | 205 | 1.953 | 1.3 | 2.933 | 0.001 |
| ever smokers | not driven | 1792 | 601 | 813 | 1.407 | 1.198 | 1.653 | <0.001 |

**Supplementary Data File D14 (continued).** Differential association within each smoking stratum.

| stratum | driven HR | not driven HR | ratio | lower | upper | p | n | deaths |
| --- | --- | --- | --- | --- | --- | --- | --- | --- |
| never smokers | 2.75 | 1.596 | 1.723 | 1.051 | 2.825 | 0.031 | 946 | 291 |
| ever smokers | 1.953 | 1.407 | 1.388 | 0.896 | 2.15 | 0.142 | 2167 | 714 |

**Supplementary Data File D15.**
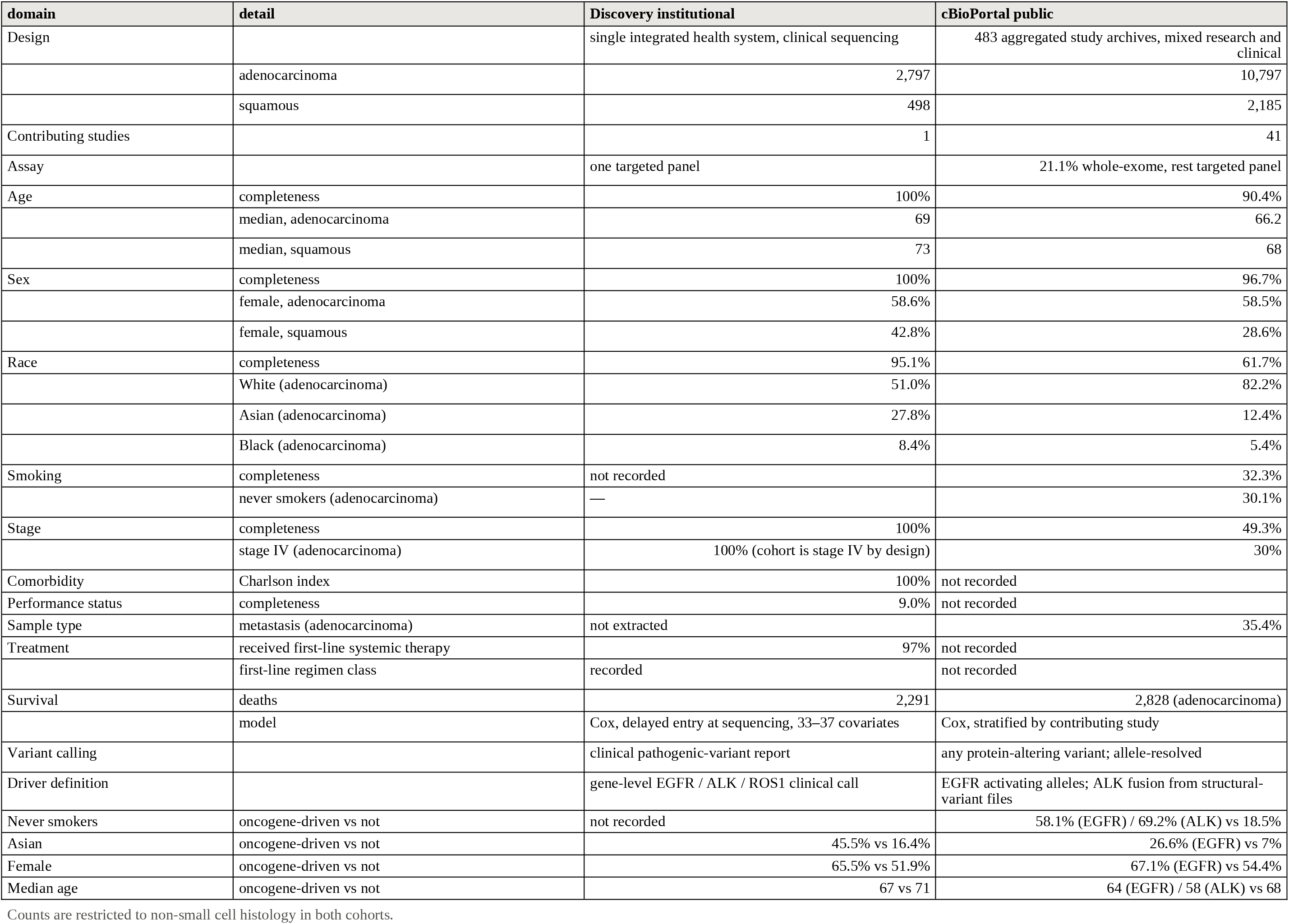
Side-by-side comparison of the two cohorts.

| domain | detail | Discovery institutional | cBioPortal public |
| --- | --- | --- | --- |
| Design |  | single integrated health system, clinical sequencing | 483 aggregated study archives, mixed research and clinical |
|  | adenocarcinoma | 2,797 | 10,797 |
|  | squamous | 498 | 2,185 |
| Contributing studies |  | 1 | 41 |
| Assay |  | one targeted panel | 21.1% whole-exome, rest targeted panel |
| Age | completeness | 100% | 90.4% |
|  | median, adenocarcinoma | 69 | 66.2 |
|  | median, squamous | 73 | 68 |
| Sex | completeness | 100% | 96.7% |
|  | female, adenocarcinoma | 58.6% | 58.5% |
|  | female, squamous | 42.8% | 28.6% |
| Race | completeness | 95.1% | 61.7% |
|  | White (adenocarcinoma) | 51.0% | 82.2% |
|  | Asian (adenocarcinoma) | 27.8% | 12.4% |
|  | Black (adenocarcinoma) | 8.4% | 5.4% |
| Smoking | completeness | not recorded | 32.3% |
|  | never smokers (adenocarcinoma) | — | 30.1% |
| Stage | completeness | 100% | 49.3% |
|  | stage IV (adenocarcinoma) | 100% (cohort is stage IV by design) | 30% |
| Comorbidity | Charlson index | 100% | not recorded |
| Performance status | completeness | 9.0% | not recorded |
| Sample type | metastasis (adenocarcinoma) | not extracted | 35.4% |
| Treatment | received first-line systemic therapy | 97% | not recorded |
|  | first-line regimen class | recorded | not recorded |
| Survival | deaths | 2,291 | 2,828 (adenocarcinoma) |
|  | model | Cox, delayed entry at sequencing, 33–37 covariates | Cox, stratified by contributing study |
| Variant calling |  | clinical pathogenic-variant report | any protein-altering variant; allele-resolved |
| Driver definition |  | gene-level EGFR / ALK / ROS1 clinical call | EGFR activating alleles; ALK fusion from structural-variant files |
| Never smokers | oncogene-driven vs not | not recorded | 58.1% (EGFR) / 69.2% (ALK) vs 18.5% |
| Asian | oncogene-driven vs not | 45.5% vs 16.4% | 26.6% (EGFR) vs 7% |
| Female | oncogene-driven vs not | 65.5% vs 51.9% | 67.1% (EGFR) vs 54.4% |
| Median age | oncogene-driven vs not | 67 vs 71 | 64 (EGFR) / 58 (ALK) vs 68 |

Counts are restricted to non-small cell histology in both cohorts.

